# SARS-CoV-2 infection dynamics in Denmark, February through October 2020: Nature of the past epidemic and how it may develop in the future

**DOI:** 10.1101/2020.11.04.20225912

**Authors:** Steen Rasmussen, Michael Skytte Petersen, Niels Høiby

**Affiliations:** Center for Fundamental Living Technology (FLinT) Department for Physics, Chemistry and Pharmacy, University of Southern Denmark; Santa Fe Institute, Santa Fe, NM 87501, USA; Center for Biosecurity and Biopreparedness (CBB), Statens Serum Institut, Denmark; Department of Clinical Microbiology, Rigshospitalet, Copenhagen, Denmark; Institute of Immunology and Microbiology, Panum Institute, University of Copenhagen, Denmark

## Abstract

**Background:** There has long been uncertainty about the relative size of the “dark” numbers, the infected population sizes and the actual fatality rate in the COVID-19 pandemic and thus how the pandemic impacts the healthcare system. As a result it was initially predicted that the COVID-19 epidemic in Denmark would overwhelm the healthcare system and thus both the diagnosis and treatment of other hospital patients were compromised for an extended period.

**Aim:** To develop a robust method for reliable estimation of the epidemic and the healthcare system load in Denmark, both retrospectively and prospectively. To do this a new pandemic simulation had to be developed that accounts for the size and the infection impact of the infectious incubating and asymptomatic infected individuals (dark numbers).

**Methods:** Our epidemic simulation is based on a SEIRS (Susceptible - Exposed - Infected - Recovered - Susceptible) model, coupled to a simple healthcare model that also includes deaths outside hospital settings. The SEIRS model has separate assessments of asymptomatic and symptomatic cases with different immunological memories. The main data used for parameter estimation in the models are hospital and *ICU* occupations, death data, serological data of antibody prevalence from the onset through August 2020 together with hospital data and clinical data about the viral infection. Optimal model parameters are in part identified by Monte Carlo based Least Square Error methods while micro-outbreaks are modeled by noise and explored in Monte Carlo simulations. Estimates for the infected population sizes are obtained by using a quasi steady state method.

**Results:** The age adjusted antibody prevalence in the general population in May 2020 was 1.37%, which yields a relative frequency of symptomatic and asymptomatic cases of 1 to 5.2. Due to the large asymptomatic population found, the actual mortality rate to date is 0.4%. However, with no behavioral and policy restrictions the COVID-19 death toll would have more than doubled the national average yearly deaths within a year. The transmission rate ℛ_0_ was 5.4 in the initial free epidemic period, 0.4 in the lock-down period and 0.8 -1.0 in the successive re-opening periods through August 2020. The estimated infected population size July 15 to August 15 was 2, 100 and 12, 200 for October 1 - 20, 2020. The efficiency of the applied daily testing strategy for both periods are estimated to be 40% of the PCR observable infected. Of more theoretical interest we demonstrate how the critical infection parameters for COVID-19 are tightly related in a so-called iso-symptomatic infection diagram.

**Conclusions:** Our simulation may be useful if a major infection wave occurs in the winter season as it could make robust estimates both for the scale of an ongoing expanding epidemic and for the expected load on the healthcare system. Our simulation may also be useful to assess a future controlled epidemic, e.g. as a basis for evaluating different testing strategies based on estimated infected population sizes. Finally, we believe our simulation can be adjusted and scaled to other regions and countries, which we illustrate with Spain and the US.

## I. INTRODUCTION

COVID-19 was a new infection that initially hit the Chinese city Wuhan. Therefore it took some time before its severity and pandemic properties were realized by China, WHO, CDC and ECDC. The Chinese outbreak was reported to WHO December 31, 2019 and its Emergency Committee declared a Public Health Emergency of International Concern January 30, 2020 and a global pandemic situation March 11, 2020 [29]. The international spread of the COVID-19 pandemic was facilitated by the Chinese New Year celebration, which took place January 15 - February 11. During this period millions of Chinese people traveled inside China and from abroad e.g. Europe and North America to China to visit their families and then returned after the end of the New Year celebration [20]. Wuhan locked down January 23, 2020 and the rest of China subsequently followed that decision [44].

Many Chinese stay and work in large European cities. One of these cities is Milan, Lombardy, Italy south of the Alps. This was the first region in a western country to be heavily hit by the pandemic officially beginning February 21, 2020 [20] [1]. In February, many schools in Europe closed for one week for winter holiday over which thousands of adults and children traveled from all over Europe to the Alps for skiing.

The first case of COVID-19 reported in Denmark came from Northern Italy February 27, 2020 and subsequently 139 Danes came home after ski-holiday in Northern Italy and Austria, mostly from Ischgl [4], where they had contracted COVID-19 during the school holiday period, which in Denmark ended February 23. The epidemic in Denmark (5.82 mil. inhabitants) developed rapidly and the Statens Serum Institut (SSI), the Danish national center for disease control, estimated that the transmission rate per infected person, ℛ_0_ was 2.6 and the prevalence increased exponentially until March 11, when WHO declared that the COVID-19 was a pandemic and Denmark closed down in the following days [9]. The adjusted information from SSI released June 11, 2020 indicates that altogether 1, 488 persons contracted the infection abroad over the first month of the Danish epidemic, initially mostly from Austria and Northern Italy. The exact number of infected that started the Danish epidemic is not clear, so after reviewing the data we decided to approximate the initial infection number to be 690 per February 24, 2020, which is also the number we use as initial conditions in our simulations [40] [17].

COVID-19 was initially imported from skiing areas in the Alps, but then local spread took place inside Denmark so the national strategy changed from prevention and containment to mitigation March 11, 2020 [40], and testing for SARS-CoV-2 was restricted to patients who needed treatment in hospitals mainly because of a severe shortage of testing equipment. After April 1st the national testing strategy was changed and more people were gradually tested leading to more information about the spread of the epidemic. This change to a more comprehensive testing strategy gradually increased and became stable between April 21 and May 17, 2020 [17].

The policies for closing down Denmark were fully implemented by March 17, 2020 and soon thereafter the number of new hospitalized COVID-19 cases decreased dramatically. The observed number of hospitalized patients already peaked April 7-8 (535 total hospital patients *H*_*tot*_, of which 146 were patients in *ICU*s). Thereafter the numbers gradually decreased.

After Easter a gradual re-opening of Denmark began April 20, 2020, and continued with fixed intervals of 2-4 weeks until June 8, 2020. Altogether, by August 31, 2020 there had been a total of 17, 084 SARS-CoV-2 positively tested persons, 3, 031 of whom were hospitalized (∼ 17.7%) and 416 of those patients were treated in *ICU*s (∼ 13.7%), while 625 infected died (∼ 3.7%). As of August 31, 2020 there were 20 hospitalized including four in *ICU*s[40].

There were five local outbreaks in Denmark between April 10 and August 31, 2020, with the last two started around August 1, 2020. One outbreak was at a meat processing plant in the city of Ringsted where the infection was introduced by a Polish worker who then mainly infected other Polish workers. Approximately 150 workers from the meat processing plant were infected before the outbreak was eliminated by mid August. The other outbreak was in the city of Århus and was associated with a soccer game July 26th and associated celebrations, a university weekend seminar attended by 80 students, the Muslim Eid festival July 30 - August 3, and a funeral of a Somalian rap-musician, which was attended by 500 people. Some of the infected unfortunately included bus drivers that spread the infection to neighboring regions. About 2/3 of the Århus cases were among people originating from Somalia and Palestine and the majority of the infected were young people. The highest incidence per 100,000 inhabitants in Århus during the outbreak was 99 and the outbreak was conquered by the end of August. [54].

Since we now have more information about the nature of COVID-19, and since there are still many unanswered questions regarding the future of the Danish epidemic, we have developed a mathematical model and a corresponding simulation of it. We calibrate the simulation using the official retrospective Danish hospital, *ICU* occupation data and death data as well as a national serological antibody test conducted late May 2020. Our simulation is used to investigate unanswered questions regarding the relative frequency of symptomatic versus asymptomatic infected, their relative infectiveness, ℛ_0_, during the pandemic, final mortality, as well as the connection to antibody development among the infected. We believe the simulation can be used as a basis for evaluating and improving contact tracing policies because it provides baselines for the infected populations sizes as well as the observable parts of these populations. Our simulation can also be used for scenario forecasting of possible future COVID-19 infection waves e.g. following a pattern similar to the well-known yearly influenza pandemic. Further we believe the model is not limited to Denmark, but is applicable elsewhere in other regions and countries dealing with the COVID-19 pandemic, if it is re-calibrated appropriately to fit the other population sizes, healthcare details and imposed policies [45] [49].

Since our model is a macroscopic description it has a number of limitations, e.g. regarding the potential impact of behaviors of individuals that agent based models would be better suited to address. As for all mathematical models, our model should be viewed as a tool to help our understanding of the current nature of the pandemic, as well as how the pandemic might develop in the future.

## II. EMPIRICAL EPIDEMIOLOGICAL DATA

### A. Main data source

To inform, test and validate our simulation we use the available data for the total hospital *H*_*tot*_ and *ICU* occupations, as well as non-hospital and total death toll numbers from the onset of the Danish epidemic through August 2020 that are published daily by SSI, Statens Serum Institut (SSI) [54], the Danish national center for disease control. The time-series of these data are shown in Fig 1.

**Figure 1:**
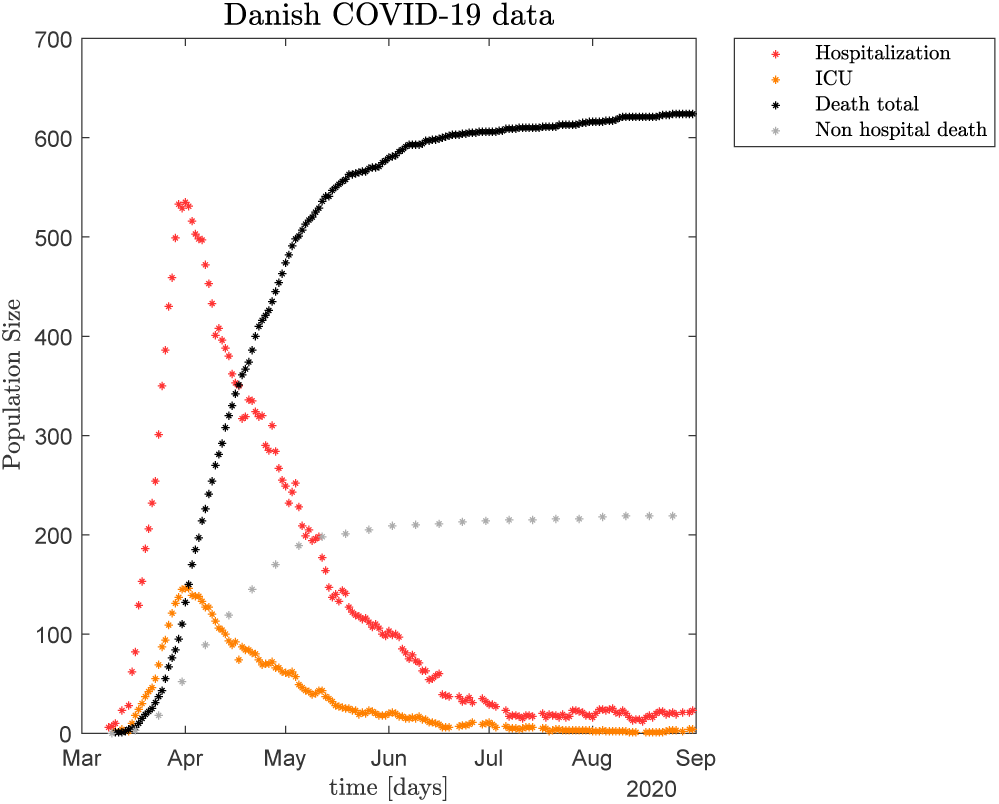
Daily total hospital *H*_*tot*_ (which includes *ICU*) and *ICU* occupation data, as well as total deaths updated daily and non-hospital (mainly eldercare facilities) deaths updated weekly from March 9 to August 31, 2020 [54].

We use additional data from SSI. These include the initial influx of newly infected individuals into Denmark in February 2020, recall discussion in previous section. We also use hospital admission data and the daily number of PCR positive individuals from both outside and inside of the hospital system, that are used to estimate the so-called daily transmission contact number, also called “contact number”, ℛ_*t*_ [54]. The ℛ_*t*_s measure the current changes in the infection trend and thus allow us to better understand the detailed (micro) properties of the empirical data after the initial infection peak.

It should be noted that SSI has modified minor details in the COVID-19 data multiple times during the period February through August 2020. However, these small changes have not had any measurable impact on our investigations or conclusions. All data are shown and further discussed in Appendix A.

### B. Asymptomatic versus symptomatic SARS-CoV-2 positive persons

Several publications have investigated the infectious potential of COVID-19 positive individuals and the ratio between asymptomatic persons and symptomatic patients. Of the 567 elderly SARS-CoV-2 positive passengers on the cruise ship Diamond Princess (median age 68 years), 46.5% were asymptomatic [35][51]. The young staff (median age 30 years) of the USS Navy carrier Theodore Roosevelt arriving in Guam was infected with COVID-19 and of the 382 surveyed service members 81.5% reported one or more symptoms whereas 18.5% were asymptomatic [31]. The passengers and staff of a cruise ship departing from Ushuaia, Argentina on a 3 week cruise to Antarctic were infected with COVID-19, of which 128 passengers and crew tested positive for COVID-19. Only 19% were symptomatic whereas 81% were asymptomatic [21]. Based on these three carefully observed groups of patients, the percentage of symptomatic persons ranges from 19 to 81.5%. The percentage of asymptomatic persons ranges from 18.5 to 81% between these 3 groups. A meta-analysis estimated that about 40% of children are asymptomatic [56]. Obviously, there is a grey zone between asymptomatic and symptomatic infected where some authors differentiate asymptomatic and weakly symptomatic while others join these two groups into one class of asymptomatic and only view more severely symptomatic as being symptomatic.

Further, SARS-CoV-2 positive persons in the last part of their incubation time, before they turn symptomatic (thus still being pre-symptomatic), have been shown to harbor the same level of virus RNA in saliva and nasopharyngeal secretions as symptomatic COVID-19 patients and therefore are assumed to be nearly as infectious as symptomatic patients [36][29][13][47][7][25]. Note that this is not the case for incubating persons that later turn asymptomatic.

The mortality of COVID-19 is calculated in two ways: A Case Fatality Rate (CFR) which is the proportion of deaths from COVID-19 compared to the total number of individuals diagnosed with the disease for a particular period. The mortality is also calculated as Infection Fatality Rate (IFR) which is the proportion of deaths among all infected individuals, including symptomatic and asymptomatic and undiagnosed subjects (e.g. based on sero-prevalence studies).

### C. Immunological memory

During COVID-19 infection an antibody response develops which involves IgM, IgG and IgA. At least 91 commercial antibody tests have been designed mostly using quantitative ELISA- or qualitative Rapid lateral flow immunoassays [42]. Nearly all patients have developed IgM, IgG and IgA antibodies 3 weeks after onset of the infection [50][55][46][8]. In one study of 70 Chinese patients, virus neutralizing antibodies correlated with antibodies measured by ELISA test [48], but in a study of 62 European patients only 66% developed protective antibodies as determined by virus neutralization tests [22]. The duration of the antibody response to SARS-CoV-2 is not yet known, but the experience from the related SARS-CoV infected patients is, that 94% of the patients still had detectable IgG antibodies after 1 year and 50-74% of the patients after 4 years [23][26]. After mild or asymptomatic MERS CoV infection, antibodies were either limited or rapidly declined within 3 months whereas the antibody response in severe infections would last for 2-3 years [23]. Preliminary results and calculations from China indicate similar dynamics of the SARS-CoV-2 antibody response [45][49].

## III. THE EPIDEMIC MODEL SYSTEM

Our basic SEIRS (Susceptible - Exposed - Infectious - Recovered - Susceptible) style model assumes a single population that can be in one of three infectious states: incubation *I*_*i*_, asymptomatic *I*_*a*_, and symptomatic *I*_*s*_. The *I*_*i*_ state is analogous to the Exposed state in the classical SEIR model although infection is also possible from the *I*_*i*_ state in our model formulation. The population starts from being susceptible *S*. After transitioning through incubation *I*_*i*_ into one of the two infectious states *I*_*a*_ and *I*_*s*_, the cases and ends up as either recovered *R* or dead *D*. All three of the infectious states *I*_*x*_, *x* = *i, s, a* can reduce the susceptible population *S* by transitioning members thereof into *I*_*i*_.

A small fraction of the recovered *R* slowly moves back to the susceptible population *S* as only a time limited immunity is assumed, which we refer to as the population’s average immunological memory. Since there likely is a significant difference in the immunological memory for the symptomatic and the asymptomatic infected we have disaggregated the recovered population into *R*_*a*_ and *R*_*s*_ with short and long immunological memory respectively. The rate of recovered population members once again becoming susceptible is given by *ξ*_*y*_*R*_*y*_, *y* = *a, s*, which is reflected in the positive terms in the first expression in equation set (1).

During the infection, a fraction of the symptomatic population *I*_*s*_ can become seriously ill and is either transferred to a hospital *H* or an *ICU*. From the regular hospital unit *H* patients can recover, move to an *ICU*, or die *D*. Patients from *ICU* can ether move to *H* or die *D*. Seriously ill individuals can also die at home or in an eldercare facility. These non-hospital deaths are indicated by *M*, where terminally ill patients arrive directly from *I*_*s*_.

A flow diagram of the infection and healthcare dynamics is shown in Fig. 2.

**Figure 2:**
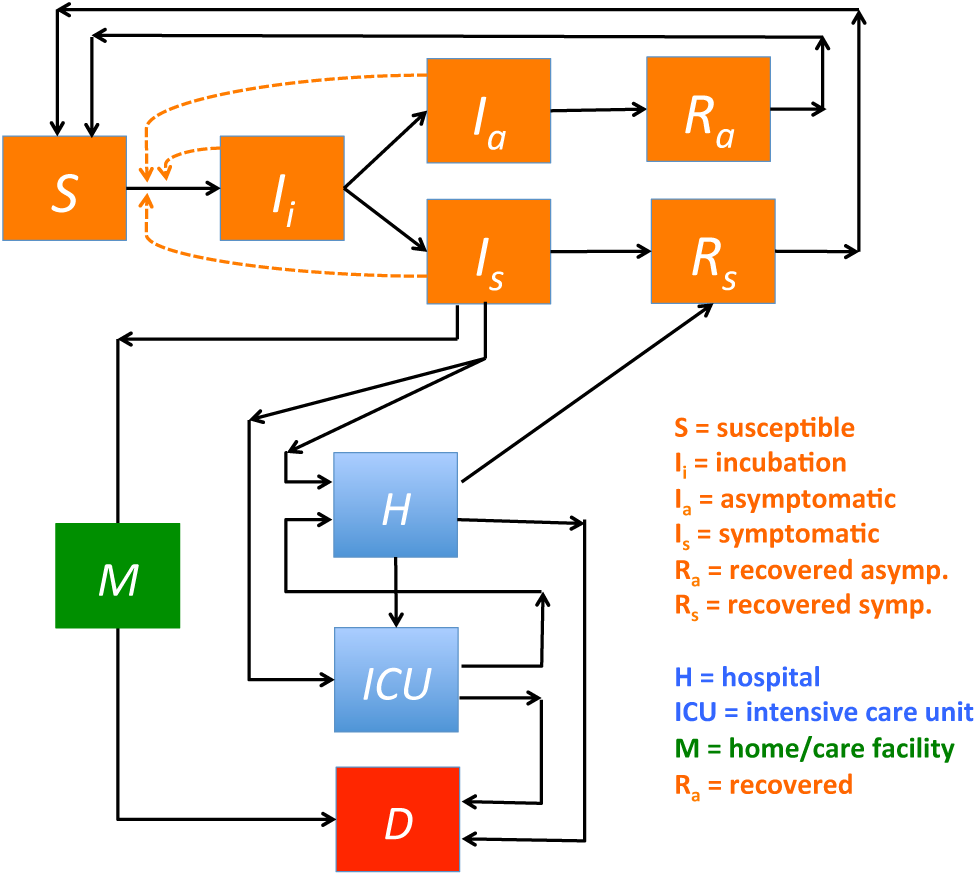
Flow through the SEIRS based model system. The equation system that defines the dynamics is given by Eqs. (1) and (2). Individuals start as susceptible *S* that can become infected and initially go through an incubation phase *I*_*i*_, then become either symptomatic *I*_*s*_ or asymptomatic *I*_*a*_, and eventually recover *R*_*s*_, *R*_*a*_ or die *D*. Note that part of the incubation phase *I*_*i*_ can also spread the infection. The dashed lines indicate the fact that the various infectious cases *I*_*x*_, *x* = *i, a, s* can infect members of the remaining *S* population. Some symptomatic infected *I*_*s*_ may become seriously ill and are either transferred to a hospital *H* or an *ICU*. From the regular hospital unit *H* patients can recover, move to an *ICU*, or die *D*. Patients from *ICU*s can ether move to *H* or die *D*. A small fraction of the symptomatic infected *I*_*s*_ symptomatic individuals may also die *D* in a non-hospital location *M*. These non-hospital deaths are also accounted for separately.

The differential equations that define the flow in Fig. 2 are given in Eqs. 1 and 2.

The epidemic dynamics is defined by SEIRS model as follows:

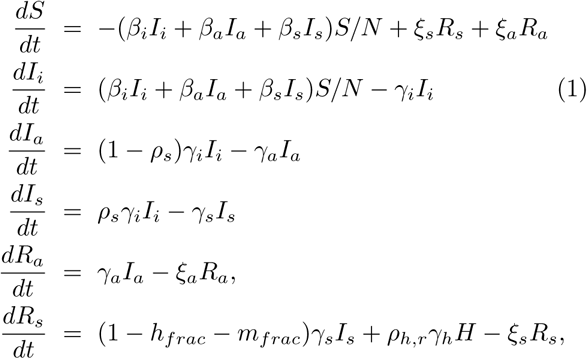

where the parameters are further discussed in Table 1. Negative terms in the above equations indicate loss of respective population members with time, while positive terms indicate population member growth. The equation for the change in the susceptible population *S* describes how susceptible are infected from three different populations *I*_*i*_, *I*_*a*_, and *I*_*s*_ with different rates *β*_*i*_, *β*_*a*_, and *β*_*s*_ normalized by the population size *N*, while recovered individuals slowly becomes susceptible again as they lose their immunity, where we assume longer immunity time 1/*ξ*_*s*_ days for the recovered symptomatic *R*_*s*_ and a shorter immunity time 1/*ξ*_*a*_ days for the recovered asymptomatic *R*_*a*_. The newly infected are moved from *S* to *I*_*i*_, the incubating population where they on average reside for 1/*γ*_*i*_ days. On average 1 − *ρ*_*s*_ of the incubated *I*_*i*_ becomes asymptomatic *I*_*a*_ where they on average reside for 1/*γ*_*a*_ days. Also, *ρ*_*s*_ of the incubated *I*_*i*_ becomes symptomatic *I*_*s*_ where they on average reside for 1/*γ*_*s*_ days.

**Table I:**
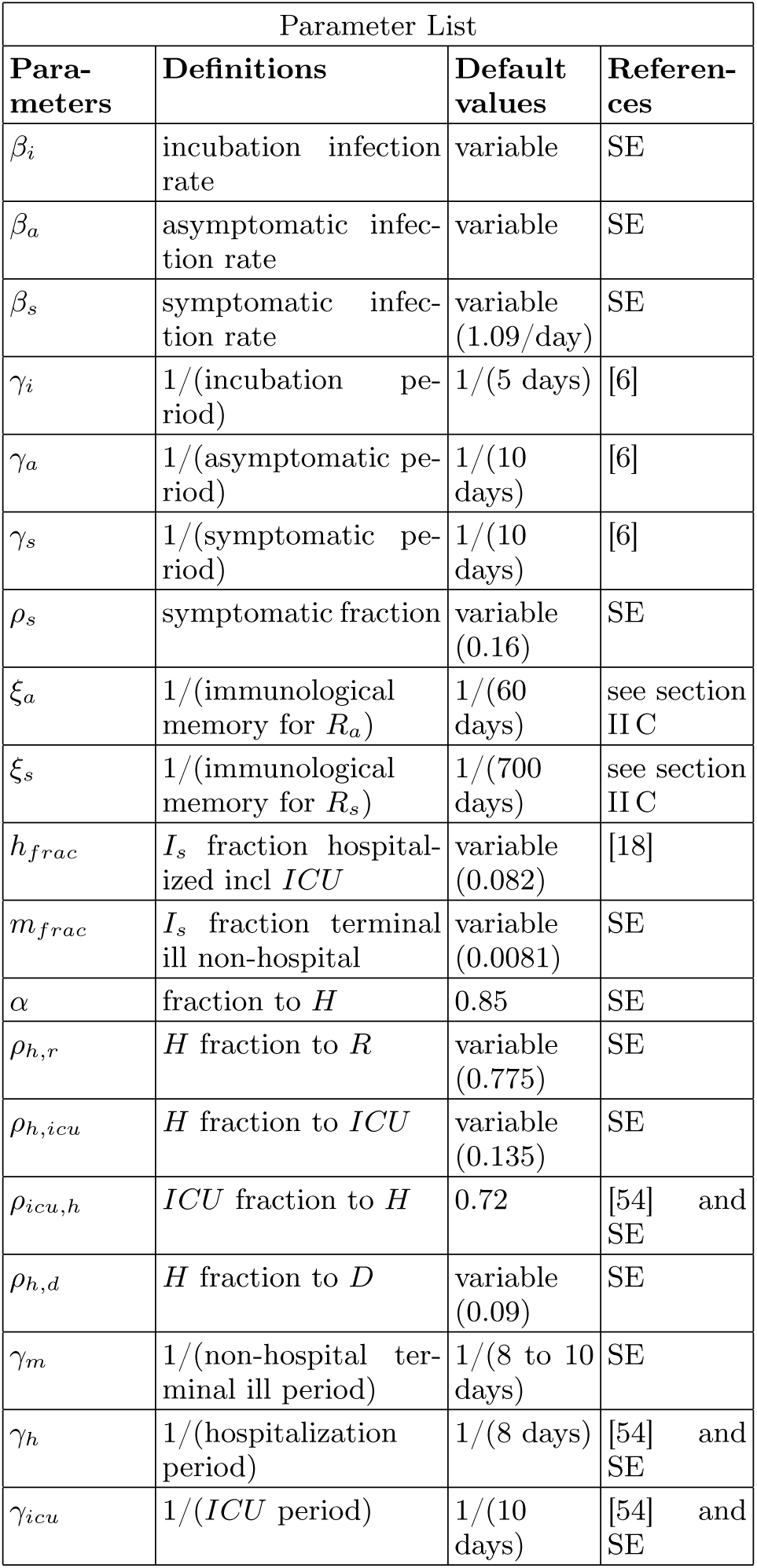
Table with input parameters to the combined SEIRS, hospital and home care model. SE = simulation estimates. The table parameter values, sometimes in parentheses, refer to a “standard run”. It will be indicated in the text if different parameters are used. Note that *ρ*_*h,r*_ + *ρ*_*h,icu*_ + *ρ*_*h,d*_ = 1.

| Parameter List |  |  |  |
| --- | --- | --- | --- |
| Parameters | Definitions | Default values | References |
| $\beta_i$ | incubation infection rate | variable | SE |
| $\beta_a$ | asymptomatic infection rate | variable | SE |
| $\beta_s$ | symptomatic infection rate | variable (1.09/day) | SE |
| $\gamma_i$ | 1/(incubation period) | 1/(5 days) | [6] |
| $\gamma_a$ | 1/(asymptomatic period) | 1/(10 days) | [6] |
| $\gamma_s$ | 1/(symptomatic period) | 1/(10 days) | [6] |
| $\rho_s$ | symptomatic fraction | variable (0.16) | SE |
| $\xi_a$ | 1/(immunological memory for $R_a$) | 1/(60 days) | see section II C |
| $\xi_s$ | 1/(immunological memory for $R_s$) | 1/(700 days) | see section II C |
| $h_{frac}$ | $I_s$ fraction hospitalized incl *ICU* | variable (0.082) | [18] |
| $m_{frac}$ | $I_s$ fraction terminal ill non-hospital | variable (0.0081) | SE |
| $\alpha$ | fraction to $H$ | 0.85 | SE |
| $\rho_{h,r}$ | $H$ fraction to $R$ | variable (0.775) | SE |
| $\rho_{h,icu}$ | $H$ fraction to *ICU* | variable (0.135) | SE |
| $\rho_{icu,h}$ | *ICU* fraction to $H$ | 0.72 | [54] and SE |
| $\rho_{h,d}$ | $H$ fraction to $D$ | variable (0.09) | SE |
| $\gamma_m$ | 1/(non-hospital terminal ill period) | 1/(8 to 10 days) | SE |
| $\gamma_h$ | 1/(hospitalization period) | 1/(8 days) | [54] and SE |
| $\gamma_{icu}$ | 1/(*ICU* period) | 1/(10 days) | [54] and SE |

However, a small fraction *h*_*frac*_ of the *I*_*s*_ population becomes severely ill and are hospitalized, see Eqs. (2), while another small fraction *m*_*frac*_ becomes so ill that they are not moved into the hospital system but stay at home or in a home care facility before dying, see Eqs. (2). All asymptomatic individuals *I*_*a*_ recover and are moved into *R*_*a*_ where we assume they retain some immunity for an average of 1/*ξ*_*a*_ days. Finally, most symptomatic infected recover *R*_*s*_ directly from *I*_*s*_, as well as from the hospital system *H*, see Eqs. (2), where we assume they retain some immunity for an average of 1/*ξ*_*s*_ days.

The dynamics of the hospital including its ICU and their deaths, as well as the non-hospital death dynamics are defined as follows:

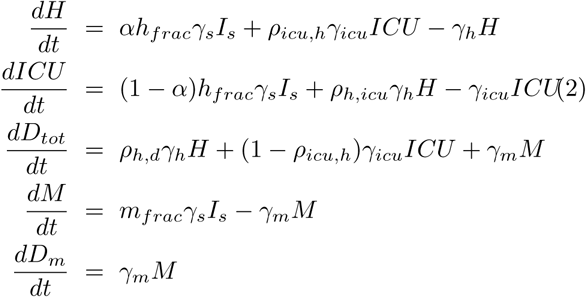

where the parameters are further discussed in Table I. Severely ill symptomatic individuals enter the hospital system at a rate given by *h*_*frac*_*γ*_*s*_*I*_*s*_ where a fraction *αh*_*frac*_*γ*_*s*_*I*_*s*_ goes directly to the regular hospital unit while (1 – *α*)*h*_*frac*_*γ*_*s*_*I*_*s*_ directly enters into an *ICU*. The size of *h*_*frac*_ is critical for the scaling of the pandemic as the hospital data are the central empirical data by which the simulation is adjusted. We use the age disaggregated data for hospital admissions from Ferguson et al. 2020 [18] together with the actual age distribution within Denmark to estimate an expected aggregated *h*_*frac*_, see Appendix A for details. The regular hospital *H* also receives a certain fraction *ρ*_*icu,h*_ of the improved patients from the *ICU* as expressed by *ρ*_*icu,h*_*γ*_*icu*_*ICU*, while patients from *H* leave the hospital after an average period of 1/*γ*_*h*_ days. Patients from *H* either recover into *R*_*s*_, or become more ill and move into *ICU*, or die and move into *D*_*tot*_. The *ICU* population, as mentioned above, receives (1 – *α*)*h*_*frac*_*γ*_*s*_*I*_*s*_ directly from *I*_*s*_ as well as *ρ*_*h,icu*_*γ*_*h*_*H* patients from *H. ICU* patients leave the population with the rate *γ*_*icu*_*ICU* and either improve and move to *H* with the already mentioned rate *ρ*_*icu,h*_*γ*_*icu*_*ICU*, or die with the rate (1 – *ρ*_*icu,h*_)*γ*_*icu*_*ICU*. The accumulated death toll stems from the hospital rate as *ρ*_*h,d*_*γ*_*h*_*H*, from the *ICU* rate as (1 – *ρ*_*icu,h*_)*γ*_*icu*_*ICU* and from the non-hospital/*ICU* associated rate as *γ*_*m*_*M*. Finally, the symptomatic population that eventually dies in a non-hospital location *M* is given by *m*_*frac*_*γ*_*s*_*I*_*s*_ with a sick period of an average 1/*γ*_*m*_ days where *m*_*frac*_ is significantly smaller than *h*_*frac*_ The non-hospital death tally are accounted for separately on a weekly basis in *D*_*m*_, while they also are counted in the total death toll *D*_*tot*_.

### A. Infection parameters and relative frequency of symptomatic and asymptomatic infected

With *ρ*_*s*_ we express the relative frequency by which an individual moves from the incubation state to the symptomatic state, while *ρ*_*a*_ = 1 – *ρ*_*s*_ expresses the frequency of movement to the asymptomatic state. We may now define the relation between the infection parameters *β*_*i*_, *β*_*a*_, *β*_*s*_ in the SEIRS model as follows:

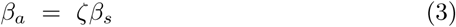

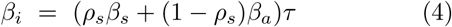

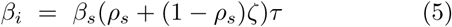

assuming that *β*_*i*_ and *β*_*a*_ can both be defined relative to *β*_*s*_ and 0 < *ζ* ≤ 1.0 since we assume the asymptomatic are always less infectious than the symptomatic infected. The value of *β*_*i*_ in Eq. (4) is defined as the weighted sum of individuals that eventually become symptomatic and asymptomatic respectively, and where *τ* may be viewed as the fraction of the incubation time they are infectious. If we e.g. assume the incubating individuals are infectious the last 1.5 days (0.3) of the average 5 day incubation time [25] we get

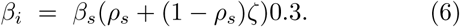

We may use clinical data to estimate a reasonable *ζ* value. Clinical investigations indicate the viral load in saliva from symptomatic and asymptomatic is comparable and that the peak viral load in saliva is found the last day of the incubation time [36] [29] [13] [47] [25]. Symptomatic children and adults have the same amount of SARS-CoV-2 in the upper respiratory tract [3].

From [28] it is further assumed that the relative infectiousness for the last day of pre-symptomatic (incubated that turns symptomatic) can be set to 1.00, while the average of the severe symptomatic, the weak symptomatic and the asymptomatic can be set to 0.89, 0.44 and 0.11 respectively during their infection time. In our study we do not distinguish between weak symptomatic and asymptomatic so the corresponding numbers for our model are 1.00, 0.89 and ((0.44+0.11)/2 = 0.275). We can now adjust Eqs. (3) and (4) so that they satisfy these relative infectiousness numbers

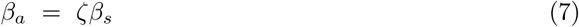

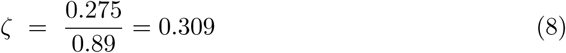

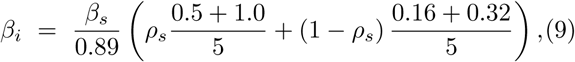

where *β*_*s*_ and *ρ*_*s*_ are defined as earlier, and we assume an incubation time of 5 days. Eqs (7) - (9) are use in the simulations. Note that Eq. (9) can be rewritten as

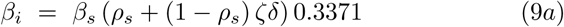

so it has the same form as Eq. (6) with the insertion of an extra factor *δ* = 1.0356 so that Eq. (9a) is identical to Eq. (9) for standard parameters, recall Table I. The main difference between this infection model and the simpler infection model given by Eqs. (3) - (6) is a slightly higher level of relative infectiousness for the incubating population for standard parameters. Simulation experiments with both types of infection models convinced us that we could obtain better fits with data if we adopt a higher weight to the incubating population. This is particularly clear when investigating the shape of the initial infection peak in the hospital data.

As neither the proportionality factor *ζ* between *β*_*s*_ and *β*_*a*_ nor the frequency *ρ*_*s*_ of symptomatic versus asymptomatic in Eqs. (4) and (9) are well known (October 2020), the initial part of this work is to explore the epidemiological impact of the relationship between *ζ, β*_*s*_, *β*_*a*_ and *β*_*i*_ as well as *ξ*_*s*_, *ξ*_*a*_ and *ρ*_*s*_ under different assumptions. Note that both infection models Eqs. (3) - (6) and Eqs. (7) - (9) assume a free epidemic. Policy interventions may be implemented by modifying the appropriate *β* parameters.

## IV. METHODS

The mathematical models defined in Eqs. (1) and (2) have a variety of parameters, where some are well defined, e.g. from clinical data, while most are restricted by the necessity for the simulations to reproduce the measured antibody data, the historical hospital and *ICU* occupation data, as well as the death data both from the hospital system and elsewhere.

The relationship between the infection parameters *β*_*i*_, *β*_*a*_, and *β*_*s*_ is based on clinical studies, recall Eqs. (3) - (6) and Eqs. (7) - (9). The value of *ρ*_*s*_, the fraction of symptomatic infected, is mainly determined by the observed serological antibody response, which was conducted May 8 - 28, 2020, and published in early June 2020 [54], together with the average antibody decay-times 1/*ξ*_*a*_, 1/*ξ*_*s*_ from the asymptomatic and symptomatic infected respectively.

The average antibody decay times 1/*ξ*_*a*_, 1/*ξ*_*s*_ are estimated based on existing knowledge about SARS and other corona viruses, recall Section II C. Despite these restrictions a few parameter combinations still have some degrees of freedom, in particular *h*_*frac*_ [18] and *m*_*frac*_ that scale the size of *I*_*s*_ and thus the whole pandemic. Also, it has been difficult to obtain explicit data for the fraction of the seriously ill that are directly admitted to a hospital *H* (*α*) versus *ICU* (1 - *α*), as well as *ρ*_*h,icu*_ and *ρ*_*h,d*_, the fraction that are moved from a hospital to an *ICU* and the fraction that dies from a hospital without going to *ICU* respectively. We will further discuss all parameters in the Results and Discussion sections.

Besides parameter variations, the mathematical models in Eqs. (1) and (2) could be made more realistic by including more details, i.e. more detailed functional relationships and more state variables. Both a geographic disaggregation and an age disaggregation would be useful, but these model extensions are outside the scope of the current study.

Our current study seeks a better understanding of the relationships between *β*_*s*_ (and thus ℛ_0_), *ζ, ρ*_*s*_ and the *ξ*_*s*_, *ξ*_*a*_ pair for the COVID-19 pandemic. It also seeks to to provide useful information about “What if?” scenarios, including potential infection wave scenarios assumed to follow the well-known winter influenza patterns. We base our investigations on the simplest model we believe is able to provide such insights. Such a model is presented in Fig. 2 and Eqs. (1) and (2). Further, by adding noise to the mean field approach defined in Eqs. and (2), we can also interpret the impact of localized micro-outbreaks of the pandemic, as well as other fluctuations found in the empirical data. We investigate the impact of noise and compare the interpretation with the mean field approach without noise.

### A. ℛ_0_ for the SEIRS model

The basic reproduction number ℛ_0_ can be derived from our SEIRS model system, recall Eqs. (1), and is given below. See Appendix D for details.

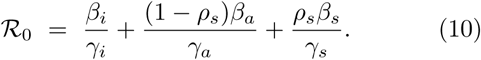

Here *β*_*x*_ is the average infection parameter for each of the infected subpopulations, *I*_*x*_, *γ*_*x*_ is one divided with the average infection period for each of the infected sub-populations and *ρ*_*s*_ is the fraction of incubating that becomes symptomatic. In the following sections we shall, when appropriate, estimate the numerical value for ℛ_0_ during the different stages of the Danish epidemic.

### B. Lockdown and re-opening of the country

The first documented COVID-19 infected arrived in Denmark in late February. On March 11, 2020 the Danish Prime minister announced that the country would be closing down in the following days, which meant that all non-essential professional activities would be shut down, social distancing (2 meters) and enhanced hand sanitation were imposed, only essential shopping was recommended (food and pharmacy), at most 10 individuals were allowed to gather at the same time, movement between different parts of the country was discouraged, international borders were closed, but face masks were not made mandatory. Further, any symptomatic individual with influenza-like symptoms were highly encouraged to self-isolate and seek medical advice and everybody who had been in contact with an infected were also highly encouraged to self-isolate.

We model the closedown process of the country as a sigmoid function for the infection parameters *β*_*s*_, *β*_*a*_, and *β*_*i*_ over a period of approximately 7 days. For all *β*_*x*_, *x* = *s, a, i*, as defined in Eqs. (3) - (9) we get

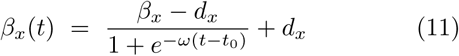

where *ω* defines the steepness of the sigmoid function, *t*_0_ the middle of the approximately 7 days lockdown process, and *d*_*s*_, *d*_*a*_, and *d*_*i*_ the infection parameters for *β*_*s*_, *β*_*a*_, and *β*_*i*_ immediately after country shut down. Once it was realized that Denmark had been hit by the COVID-19 pandemic, all individuals with flu-like symptoms were highly encoraged to self-isolate, so in the following we assume *β*_*s*_ remains at the low level ∼ 1% of the free pandemic value, both during lockdown and after the country reopens.

We model the reopening of the country in a manner similar to the closedown process, i.e. by increasing *β*_*a*_ and *β*_*i*_ following the dates where the national reopening policies change. The reopening starts April 20 with the reopening of schools for the youngest kids (K-5) together with a number of small businesses including dentists, hairdressers and other businesses where services are rendered and where close physical proximity between a provider and a costumer is necessary. Later reopening activities were implemented May 7, 20, June 8, and August 14, 2020 [33].

We chose to model the reopening process as a slow (linear) increase of *β*_*a*_ and *β*_*i*_ over that interval. Thus, at time *date*_1_ April 20, 2020, when the country starts reopening activities to *date*_2_, June 8, 2020, a slow linear increase in *β*_*y*_, *y* = *i, a* occurs as defined below:

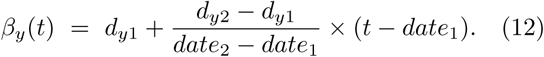

At time *date*_2_, June 8, 2020, this linear increase levels off and becomes flat with a constant 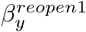 value. For the second reopening, August 14 - 31, 2020, a linear increase is again similarly defined leading to a constant 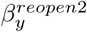 using the August 31, 2020 value.

A graphical depiction of the reopening dynamics as expressed in Eqs. (11) and (12) is shown in Fig. 3.

**Figure 3:**
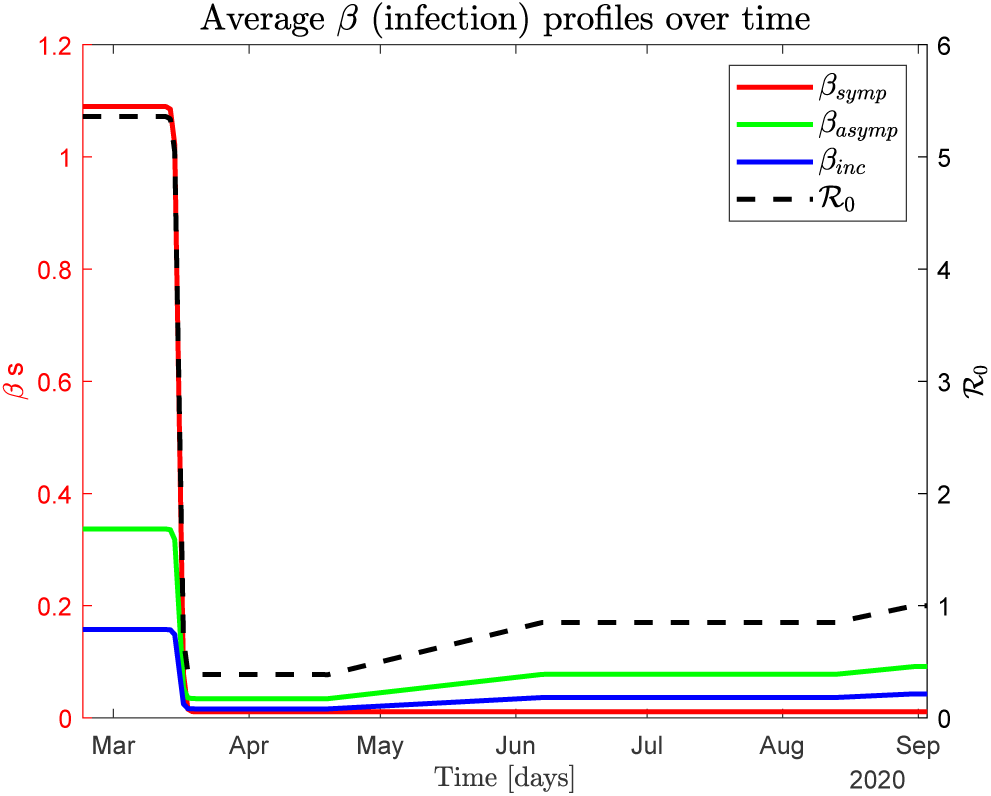
The middle of the governmental lockdown process of the country occurs approximately March 16, 2020, 20 days after the arrival of the initial group of COVID-19 infected. The lockdown is modeled by a sigmoid function with the inflection point at the middle of the process, see Eq. (11) and text for details. Successive governmental interventions that gradually reopen the country are approximated as a linear ramp starting April 20 and leveling off to a constant level June 8, 2020. The last reopening, August 14 - 31, 2020, is handled similarly, see Eqs. (11) and (12) and text for details.

### C. Monte Carlo optimization of parameters

We may adjust the *β* parameters in Eqs. (1) and (2) manually while at the same time ensuring that the scale of the infection fits the data by adjusting *ρ*_*s*_ that determines the relative sizes of the symptomatic and asymptomatic populations. From the national serological test conducted in May 2020, the estimated number of recovered individuals is reported to be about 1.3% of the population (May 28, 2020) although this study only includes individuals of age 18 or above.

The advantage of such an approach is that after some time experimenting with the simulation, the experimenter obtains an intuitive understanding of how the simulation reacts to parameter changes and minor parameter adjustments can be made iteratively.

Alternatively, we may use a Monte Carlo method (MC) to randomly choose the parameter combinations from a uniform distribution in given intervals in a given scenario. This is done until a desired minimum value of the sum of squared differences (LS) between the daily reported data for total Danish hospital occupations and the same data generated in the corresponding simulation is achieved. For more details, see Appendix C.

Using the MC-LS method we initially need to ensure the optimization process takes into account the appropriate scale of the outbreak that can be obtained by adjusting *ρ*_*s*_. Thus we start the formal parameter optimization by initially looping over slightly different *ρ*_*s*_ and different 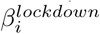 and 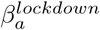 values followed by a two-step MC-LS process:

i. In the first step we optimize the *β*^0^’s at the onset of the outbreak using the infection model given in Eqs. (7) - (9) and optimize for the period February 24 - April 20, 2020. We assume the lockdown 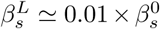 of the outbreak value once the lockdown in a place, as virtually all symptomatic individuals are isolated. We can now optimize for 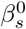 (as well as 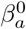 and 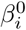) in the free outbreak and lockdown period from February 24 to April 20 for different fixed 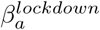 (and 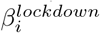) values, where each optimization yields a slightly different 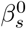: The lower 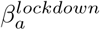 (and 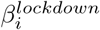), the higher 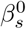.
ii. In the second step we optimize two parameters simultaneously, 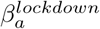 and 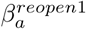 (as well as 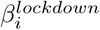 nd 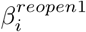) from April 20 to August 14 with the corresponding 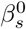 obtained in step (i), also using the infection model given in Eqs. (7) - (9).

Going through the simulation with correct seroprevalence numbers for May 28, 2020, the MC solutions with the smallest LS error in the second step is then selected. Then the output from the MC-LS optimization yields the following set of five parameters: 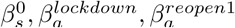 and thus the associated 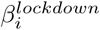 and 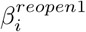.

As we shall see in the Results Section, the MC-LS method tends to slightly underestimate 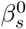 compared to a visual inspection mainly because of the historical details of the data that are discussed in the next subsection.

### D. The impact of noise

It is somewhat problematic using our mean field modeling approach to analyze and understand the successive, small and localized epidemic outbreaks over the late spring and summer of 2020. Although these outbreaks are small due to the low national hospital occupation numbers from May through August, most of these later micro-outbreaks are actually reflected in the hospital admittance numbers. Further, as we inspect the hospital admittance numbers for mid April 2020 they reveal a “shoulder” in the data that can be seen as one or more larger localized outbreaks overlaying a steadily decreasing macroscopic trend in the hospital admittance data, which is most clearly seen in Fig. 13(2), Appendix A. Both these earlier (larger) and later (smaller) fluctuations in the hospital data are also picked up in SSI’s daily estimated “contact number” ℛ_*t*_ that essentially measures the deviation from the currently observed number of infected compared to an average over that last week [54]. ℛ_*t*_ < 1 indicates decreasing observed infected, ℛ_*t*_ > 1 indicates increasing observed infected, while ℛ_*t*_ ∼ 1 means no change in the measured infection level. ℛ_*t*_ is estimated both from hospital admittance data and from the observed infected (positively PCR tested) individuals in the population at large.

When minimizing the least square error between the generated total hospital occupation *H* + *ICU* in simulation and the reported total hospital occupation, we obscure these fluctuations in the observed data, which is unsatisfactory as we also hope to understand the smaller details in the pandemic, in particular the micro-outbreaks.

This issue can be circumvented by viewing the localized outbreaks as events generated by coincidences that can be implemented by adding a noise term to the equation set (1) that determines the dynamics for *I*_*i*_ as follows:

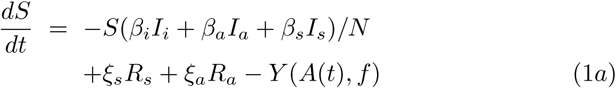

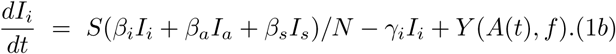

Here *Y* (*A*(*t*), *f*) is a Poisson point process with frequency *f* and amplitude *A*(*t*), a local outbreak of size *A* at time *t* that on average occurs with frequency *f*. We have included –*Y* (*A*(*t*), *f*) in Eq. (1a) to maintain population conservation by balancing out the same term in Eq. (1b). The rest of the Eqs. (1) and (2) remain the same.

In the Results Section we review manual parameter adjustment with and without noise as well as the MC-LS parameter optimization without noise.

### E. Quasi steady state approximation

For extended periods the hospital occupation may be approximately constant indicating a steady state situation of the pandemic. For example, we saw that during the period July - August, 2020. If we assume a quasi steady-state pandemic situation we approximately have:

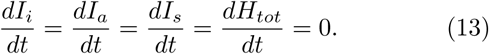

We can then use our model to estimate the sizes of the infected background populations *I*_*i*_, *I*_*a*_, and *I*_*s*_ as we can calculate backwards from a known approximately constant hospital occupation. In a steady state situation we have *h*_*frac*_*γ*_*s*_*I*_*s*_ − (1 − *ρ*_*h,icu*_)*γ*_*h*_*H* − (1 − *ρ*_*icu,h*_)*γ*_*icu*_*ICU* = 0 that expresses: what goes into the hospital system equals what leaves the hospital system. With known parameters *h*_*frac*_, *γ*_*s*_, *ρ*_*h,icu*_, *γ*_*h*_ and known *H* and *ICU* populations the size of *I*_*s*_ can be estimated. Then *I*_*a*_ = ((1 − *ρ*_*s*_)/*ρ*_*s*_)*I*_*s*_ and *I*_*i*_ = (*I*_*a*_ +*I*_*s*_)/2 as the average incubation time is half the average time of both the symptomatic and asymptomatic.

## V. RESULTS

### A. Visual inspection for infection parameter fitting

The typical infection dynamics generated by the SEIRS model as defined by the equation system (1) are discussed in Fig. 4 while the corresponding hospital and non-hospital dynamics defined by equation systems (2) are discussed in Fig. 5. For this simulation, the fraction of symptomatic infected is *ρ*_*s*_ = 0.16 and the infection model is defined by Eqs. (7) - (9), where 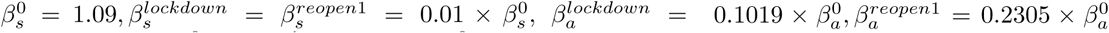. These parameters are selected by visual inspection of simulation output ensuring an infection scale such that ∼ 79,700 individuals had been infected by May 28, 2020 [54]. Other parameter values used are shown in Table I.

**Figure 4:**
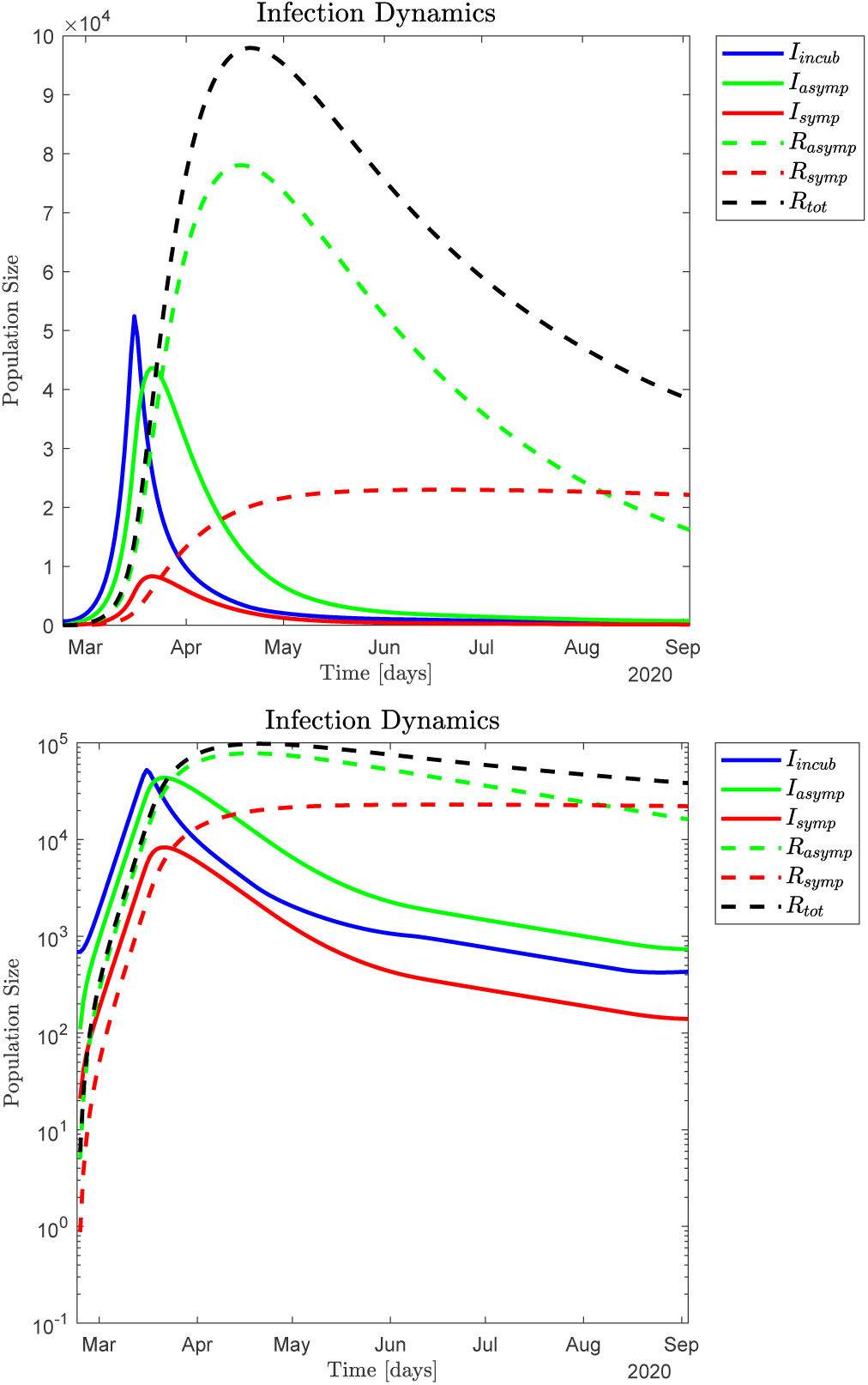
Infection dynamics; *upper panel* linear scale, *lower panel* logarithmic scale. We assume the initial infection *I*_*i*_ stems from 690 individuals entering Denmark February 24, 2020. Parameter adjustments were done by visual inspection to match the observed hospital and death data in Fig. 5, as well as the number of persons with COVID-19 antibodies ∼ 1.37% of the population, ∼ 79, 700 recovered on May 28, 2020 (day 95 of the simulation). The governmental interventions, a lockdown and a successive reopening of the country, are discussed in Fig. 3. In this simulation *ρ*_*s*_ = 0.16, which means that a little more than 5 out of 6 infected individuals end up being asymptomatic, and we use the relationships between the *β*’s given in Eqs. (7) - (9). 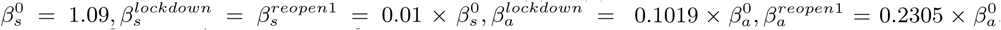. The immunity loss parameters for asymptomatic infected are 1/*ξ*_*a*_ = 60 days and for symptomatic infected 1/*ξ*_*s*_ = 700 days. Health care parameters: *α* = 0.85, *γ*_*h*_ = 1/8, *ρ*_*h,r*_ = 0.775, *ρ*_*h,icu*_ = 0.135, *ρ*_*h,d*_ = 0.09, *γ*_*icu*_ = 1/10, *ρ*_*icu,h*_ = 0.72, *m*_*frac*_ = 0.0081, *γ*_*m*_ = 1/8. These are the standard parameters shown in Table I. The central relationship between *ζ, ρ*_*s*_, *β*_*s*_ and ℛ_0_ is discussed in Fig. 8.

**Figure 5:**
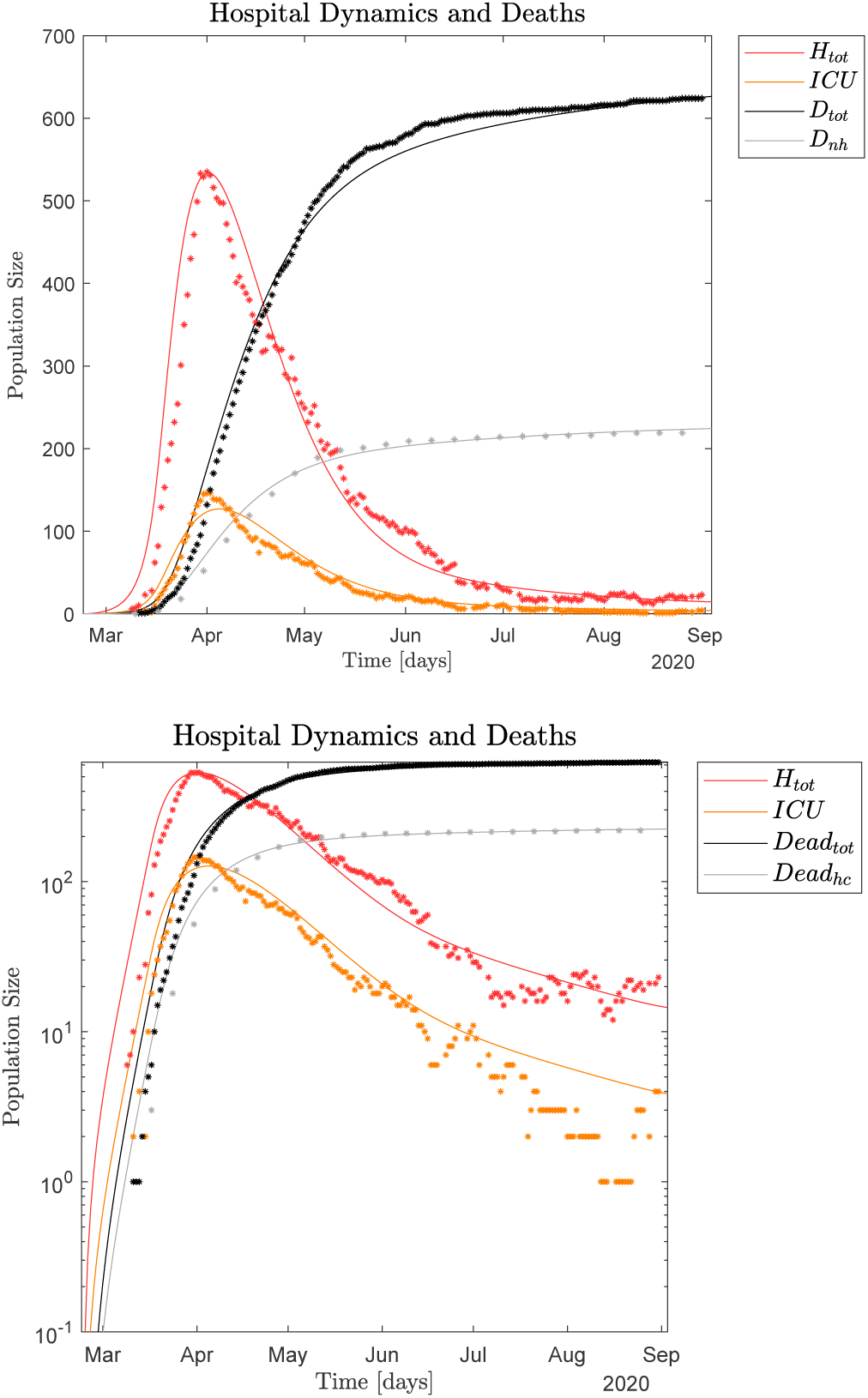
Dynamics of total daily hospital occupation *H*_*tot*_, *ICU* occupation, total death *D*_*tot*_ as well as weekly non-hospital deaths *D*_*nh*_ corresponding to (same simulation as) infection dynamics in Fig. 4; *upper panel* linear scale; *lower panel* logarithmic scale. Dots represent the observed hospital and death data, lines represent the simulation data. Parameter adjustments were done by visual inspection to match observed data. Health care parameters used are *α* = 0.85, *γ*_*h*_ = 1/8, *ρ*_*h,r*_ = 0.775, *ρ*_*h,icu*_ = 0.135, *ρ*_*h,d*_ = 0.09, *γ*_*icu*_ = 1/10, *ρ*_*icu,h*_ = 0.72, *m*_*frac*_ = 0.0081, *γ*_*m*_ = 1/8, which are the standard parameters discussed in Table 1. These hospital and death times series data together with the hospital parameters define our main means of verification (or falsification) of our simulations. The central relationship between *ζ, ρ*_*s*_, *β*_*s*_ and ℛ _0_ is discussed in Fig. 8.

It is noted that the large size of the infected total used above is because we also include the asymptomatic infected, which are estimated to be ∼ 5.2 times larger than the symptomatic infected. Initially only the symptomatic were tested and this is still reflected in the verified positive PCR test numbers that are commonly quoted.

### B. Monte Carlo Least Squares optimization of infection parameters

Alternatively, we can utilize a Monte Carlo based Least Squares (MC-LS) optimization method to identify the different *β*_*x*_ where *x* = *s, a, i* as discussed in Section IV C. The result of the simulation is shown in Fig. 6. The MC-LS parameter optimization (8, 000 iterations) yields (assuming 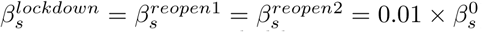), *ρ*_*s*_ = 0.1570 and 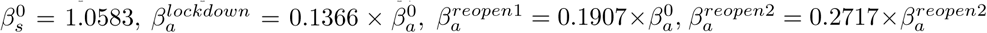 using the same infection model as above defined by Eqs. (7) - (9). Also the death rates have to be slightly increased to fit the data, see discussion in Fig. 6. Other parameter values used are shown in Table I, except the parameters controlling the death rates both from the hospitals and the non-hospital locations.

**Figure 6:**
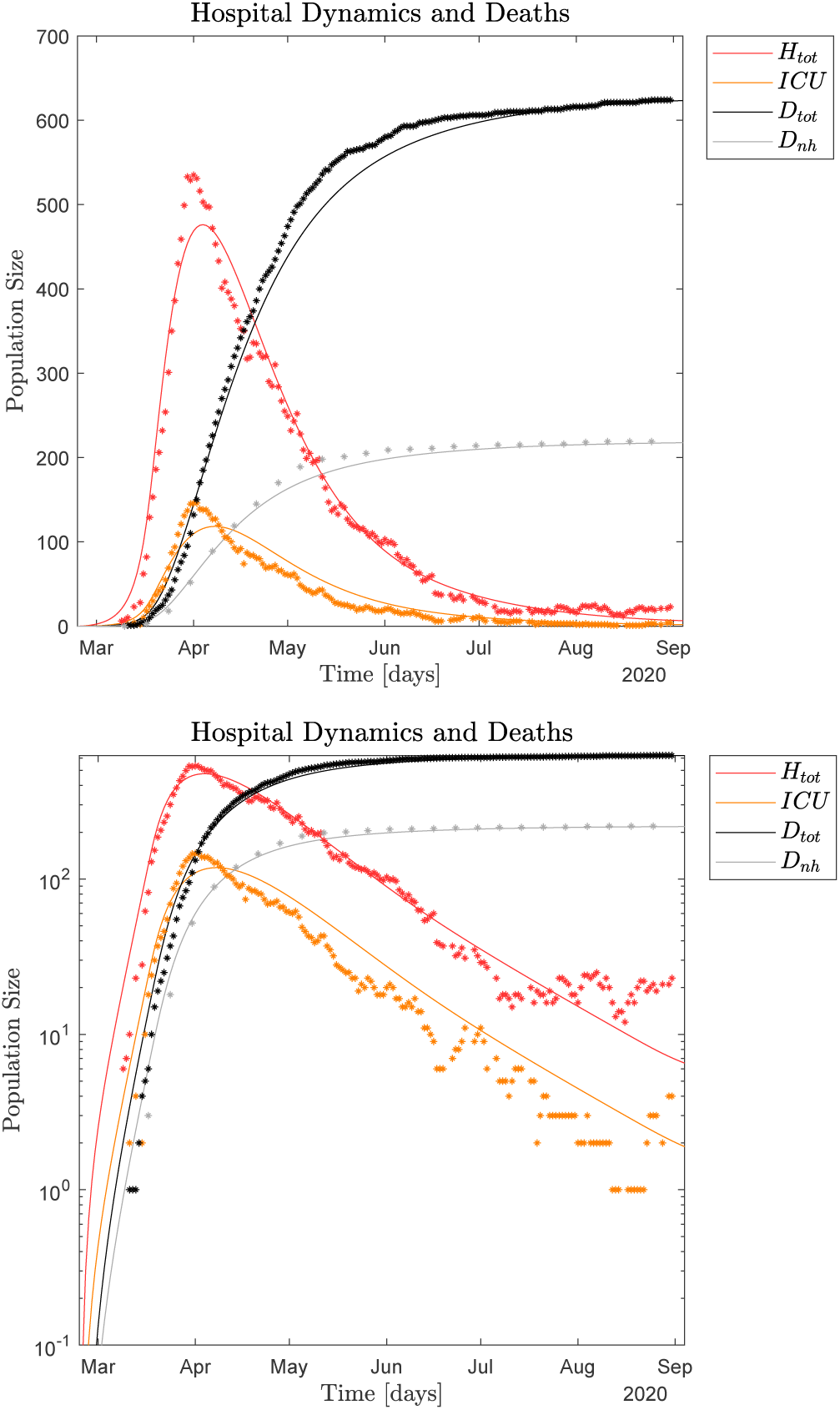
Dynamics of total daily hospital occupation *H*_*tot*_, *ICU* occupation and total death *D*_*tot*_ dynamics as well as weekly non-hospital deaths *D*_*nh*_, where the parameter adjustments are done by 8, 000 iterations of MC-LS optimization. *Upper panel* linear scale and *lower panel* logarithmic scale. Dots represent the observed hospital and death data, lines represent the simulation data. See text for parameter details and compare with Fig. 5. Note how the MC-LS algorithm optimized for best fit of *H*_*tot*_ chooses a slightly lower 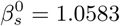 because it minimizes the sum of the squares of the errors, which means that it misses the full amplitude of initial infection peak. A slightly lower 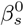 implies a slightly lower *ρ*_*s*_ = 0.157 (relatively more asymptomatic infected needed to reach the measured seroprevalence level per May 28, 2020). Further, a lower 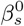 implies a higher 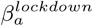 level (March 17 - April 20, 2020) and a relatively higher 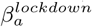 level implies a relatively lower 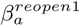 level (after June 8 and before August 15, 2020). Finally, a lower 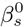 requires a slightly higher death rates to match reported data so *ρ*_*h,r*_ = 0.775 → 0.755; *ρ*_*h,icu*_ = 0.135 → 0.15; *ρ*_*h,d*_ = 0.09 → 0.095; and *m*_*frac*_ = 0.0081 → 0.0083.

The hospital occupation data has a “shoulder” after the peak (see Appendix A and Fig. 13(2)). This means there is an extended period in mid April 2020 with close to constant hospital admissions. Thus the MC-LS algorithm selects a 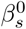 that underestimates the observed impact on the health care system as the LS error becomes smallest if the simulated hospital curve is close to as many observed data points as possible. Therefore the MC-LS estimate misses the full scale of the hospital occupation peak. As a consequence of the initial underestimation of 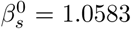, the MC-LS algorithm appropriately compensates by slightly increasing the estimate for 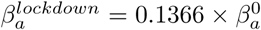, again to minimize the least square error between reported and generated data. Compare Figs. 5 and 6.

### C. Impact of noise

Yet another way to interpret the Danish COVID-19 pandemic data is to view the macroscopic infection dynamics as described by the SEIRS model and the microscopic events as superimposed noise. Thus the noise can be interpreted as microscopic, localized infection events, as discussed in Section IV D.

We may assume the impact of noise in the hospital data only becomes visible around April 1, as singular microscopic events would be difficult to distinguish earlier because of the dominating macroscopic growth of the pandemic given by 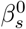 and the successive macroscopic disruption of the infection dynamics due to the sudden lockdown of the whole country. Inspecting the data from April 1 to August 31 supported by ℛ_*t*_ [54] measured from both hospital admissions and observed infected numbers, yields five distinct peaks corresponding to microscopic events, see Fig. 13(5 - 6) Appendix A. Thus we may estimate *f* as 5 events/150 days = 0.033 events/day.

Now recall Eqs. (1a) and (1b). For April 1 - 20, 2020, we assume *A*(*t*) = 2, 500 based on the size of shoulder in the hospital admittance for mid to late April; recall Fig. 13(5 - 6). *A*(*t*) is thus a bit smaller than 10% of the average size of the total infected population *I*(*t*) = *I*_*i*_ +*I*_*a*_ +*I*_*s*_ in that time interval according to our simulations. For April 21 - August 31, 2020, we assume *A*(*t*) = 1, 000 as there are no more major “shoulders” in the data. Thereby *A*(*t*) becomes approximately 1/3 of the size of the total infected population from early July through August, 2020 according to our simulations.

As a basis for the noise added simulation we in part use the standard parameters previously used to generate Figs. 4 and 5: *ρ*_*s*_ = 0.16, 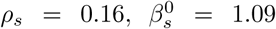 and 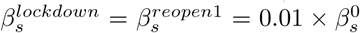. However, the reopening parameters have to be lower 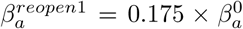 and 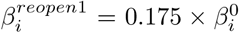. These *β* values are lower so that they, together with the noise added infections, on average generate a similar total number of infections as in the reported data. For simplicity we do not include the last reopening August 14 - 31, 2020, as it does not impact the reported data. Fig. 7 shows a comparison between the observed data and data generated by 100 Monte Carlo simulations each with a different noise sequence realization. Two such noise induced simulations are also highlighted in Fig. 7 to give an impression of the trajectories of individual realizations. Compare Fig. 7 with Figs. 5 and 6.

**Figure 7:**
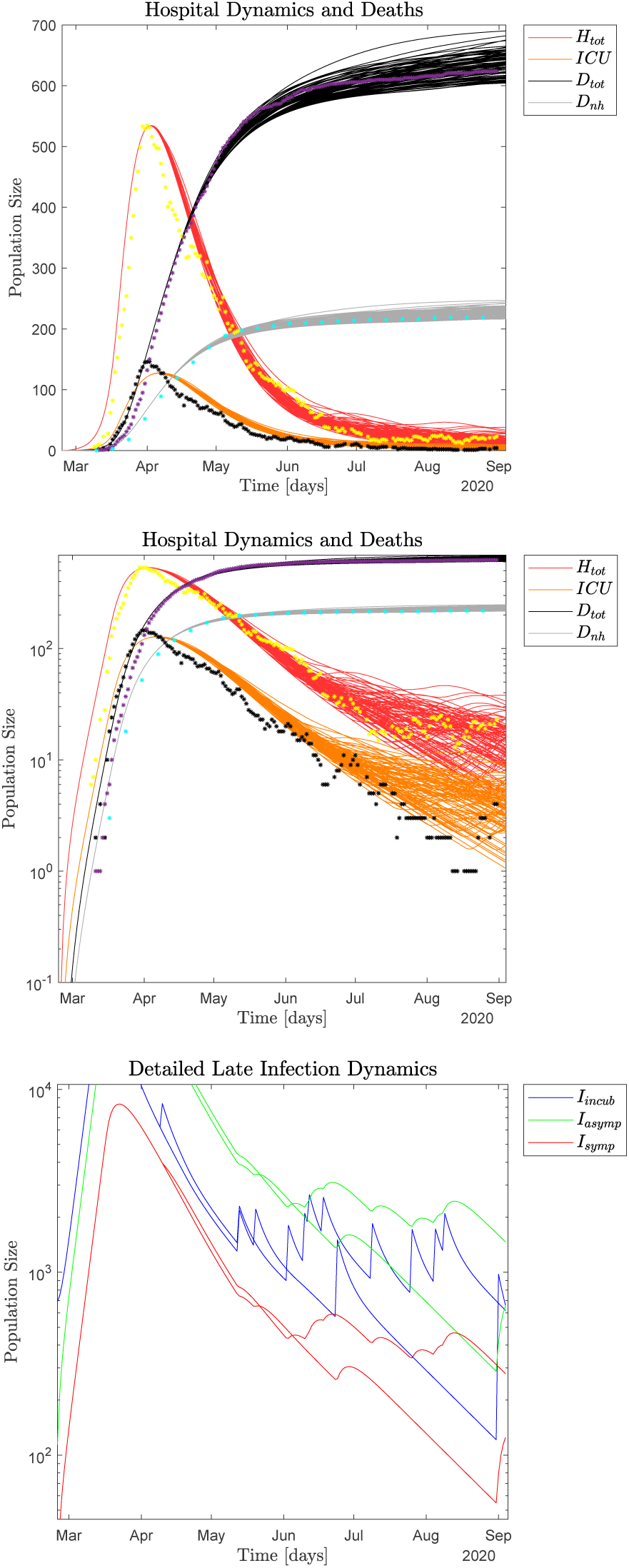
100 Monte Carlo realizations of the epidemic with noise added micro-outbreaks with a frequency of 0.033 events/day and an amplitude of size *A*(*t*) = 2,500 for April 1 - 20, and 1, 000 after April 21, 2020. *Upper panel*: linear scale dynamics of total daily hospital *H*_*tot*_ and *ICU* occupations together with total daily deaths *D*_*tot*_ as well as weekly non-hospital deaths *Dnh*. Dots represent the observed hospital and death data, lines represent the simulation data. *Middle panel*: logarithmic scale dynamics of the same observables. *Lower panel*: two realizations of the noise induced impact of local outbreaks on *I*_*i*_(*t*), *I*_*a*_(*t*), *I*_*s*_(*t*) (logarithmic scale). See text for details.

### VI. SYMPTOMATIC VERSUS ASYMPTOMATIC POPULATIONS

In the current study we define the symptomatic infected population as consisting of individuals with serious symptoms, while the asymptomatic population for simplicity includes individuals with no symptoms as well as weak symptoms, e.g. minor symptoms from upper airways (nose, throat) and no fever. Thus, in our model a symptomatic infected individual knows that he or she is sick and will likely seek medical advice as well as self-isolate, while an asymptomatic individual likely would continue his or her daily routines at home, go to school or go to work as well as engage in social activities. In some model studies asymptomatic and weakly symptomatic are treated separately [28].

Because historical total hospital *H*_*tot*_ and *ICU* occupations as well as death data only stem from the symptomatic infected population *I*_*s*_, and we assume a constant fraction of seriously ill individuals are in need of hospitalization [18], the symptomatic population size is at any given time constrained by these real life data. The same is true about the non-hospital deaths. In contrast, the asymptomatic population could in principle vary freely if we didn’t include additional empirical information.

Randomized serological tests for SARS-CoV-2 antibodies in a population yields an estimate of the recovered, both symptomatic and asymptomatic, individuals at any give time. These recovery numbers can then be compared to the recovered population generated in simulation. The size of the recovered populations at any given time depends on both the number of recovered infected individuals and on the decay rate of the average immunological memory of symptomatic *ξ*_*s*_ and asymptomatic *ξ*_*a*_ individuals, recall Eqs. (1). This means that with only one serological measurement at *t*_1_ infinitely many combinations of *ρ*_*s*_ and *ξ*_*s*_, *ξ*_*a*_ fit the observed serological prevalence. In principle we need two serological samples at times *t*_1_ and *t*_2_, with some months in between, to fix the parameter values for *ρ*_*s*_ and the average of the two *ξ*’s. Thus, measuring the recovered population at two different times *t*_1_ and *t*_2_ fixes the average *ξ*, which then can be expressed in days.

Alternatively, we can estimate the immunological memory based on our knowledge of other corona viruses, recall Section II C, and use these as parameter inputs to our simulations. Assuming a half-life for 1/*ξ*_*s*_ to be 700 days and 1/*ξ*_*a*_ to be 60 days respectively, we can determine *ρ*_*s*_ and thus the relative frequency of symptomatic versus asymptomatic individuals from only one randomized serological test at time *t*_1_ [49].

The Danish SARS-CoV-2 seroprevalence was found to be 34 out of 2, 424 tested ∼ 1.4% of the Danish population during the period May 8 - 28, 2020 [54], where it should be noted that this seroprevalence study only includes adults of more than 18 years of age. However, about 1/3 of the Danish population has been PCR-tested for SARS-CoV-2 virus by the end of August 2020 and the infection prevalence was found to be ∼ 1.1% in adults and ∼ 0.6% in children [54]. Since there are ∼ 1.30 mil. 0-19 aged and ∼ 4.52 mil. 20-90+ aged in Denmark we estimate 0.6 × (1.30/5.82)×1.4%+1.1 × (4.52/5.82) × 1.4% = 1.3692% ∼ 1.37%, where we assume the age distribution of infection prevalence is similar to the age distribution in seroprevalence.

We now need to adjust *ρ*_*s*_ to obtain ∼ 0.0137 × 5.82 × 10^6^ 79, ∼ 790 recovered individuals in our simulation by the end of May, 2020. This yields a *ρ*_*s*_ *≃* 0.16, which means that we estimate that approximately 16% of the infected are symptomatic while 84% of the infected are asymptomatic; thus ∼ 5.2 times more asymptomatic than symptomatic infected individuals. Also, as Denmark only has conducted one prevalence study so far, our estimated scale of the pandemic significantly relys on this one measurement.

### A. Iso-symptomatic infection diagram

The parameters *ζ*: the relative infectiousness of asymptomatic versus symptomatic; *ρ*_*s*_: the relative fraction of infected (incubated) that becomes symptomatic; 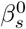: the infection parameter; and the initial ℛ_0_, define critical properties of COVID-19 and other communicable infectious diseases. In Fig. 8 shows the relationship that exists between 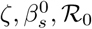, and 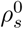, which we may call an *iso-symptomatic infection diagram*. We assume *ζ, ρ*_*s*_, *β*_*s*_ are constant over time and throughout a free (non-restricted) pandemic. Thus 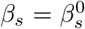 and also the initial ℛ_0_ are defined for the initial free pandemic period.

**Figure 8:**
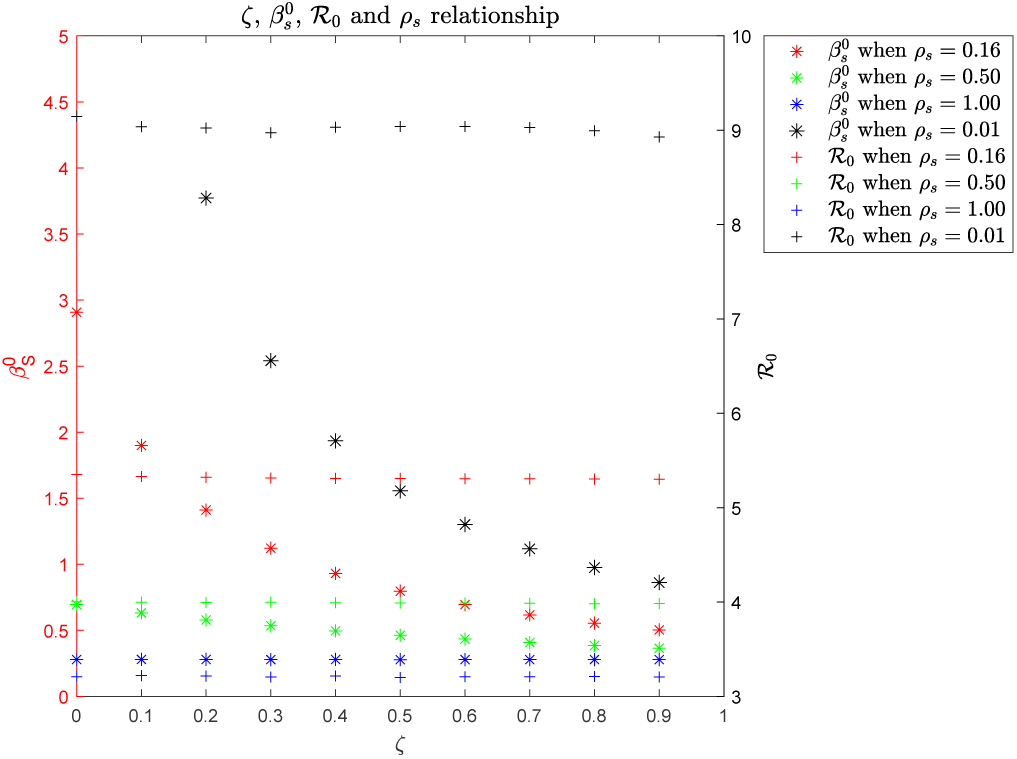
The *iso-symptomatic infection diagram* shows the characteristics of a free COVID-19 epidemic as when it began in Denmark and is constructed as a result of multiple MC-LS optimization simulations for the free pandemic using the infection model in Eqs. (3) - (6). The abscissa, the *ζ* parameter, defines the relative infectiousness between symptomatic and asymptomatic infected (*β*_*a*_ = *ζβ*_*s*_). The right ordinate is the initial reproductive number ℛ_0_ (the number of persons which are infected by one contageous person) and the “+” curves indicate how it depend on *ζ* for *ρ*_*s*_ = 1.0, 0.5, 0.15, 0.01, the relative frequency of symptomatic *ρ*_*s*_ versus asymptomatic (1 – *ρ*_*s*_) infected. Note how these “+” curves are virtually horizontal, see text for details. The right ordinate starts with ℛ_0_ = 3.0 so that it can match the left ordinate depicting the infection rate 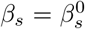 of symptomatic infective individuals at the beginning of the epidemic. Different *ρ*_*s*_ values give different families of “* “curves for *β*_*s*_ where the *ρ*_*s*_ value colors match the *ρ*_*s*_ colors for the corresponding “+” curves for ℛ_0_. Note that *β*_*s*_ as a function of *ζ* for *ρ*_*s*_ = 0.01 is off the chart (i.e. *β*_*s*_ ≃79.5) and also *ρ*_*s*_ → 0 implies *β*_*s*_ → ∞. The best parameter combination for describing the Danish COVID-19 pandemic with the infection model given in Eqs. (3) - (6) is discussed in the text. Note that an often cited ℛ_0_ for COVID-19 is ∼ 3.2, which is obtained if no distinction is made between the symptomatic and asymptomatic infected populations: *ζ* = 1.0 and *ρ*_*s*_ = 1.0.

The iso-symptomatic infection diagram is constructed from multiple Monte Carlo - Least Square parameter optimized simulations that *all* match the observed Danish COVID-19 hospital occupation data at the onset of the pandemic. In this construction we use the more generic infection model with variable *ζ* as defined in Eqs. (3) - (6).

Quantitatively almost identical iso-symptomatic infection diagrams are found when we optimize the involve parameters not only for the initial free pandemic, but also include data through April 19 (lockdown) or include data through August 14, 2020 (after the first reopening).

Note that in Fig. 8 the “+” curves for ℛ_0_ are virtually horizontal, which means that *ζ* and 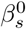 balance each other out; as *ζ* increases 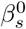 decreases and vice versa. This can be seen by substituting *ζ* into the expression of ℛ_0_ in Eq. (10) that after reducing the expression yields 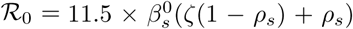 where we have also inserted *γ*_*a*_ = *γ*_*s*_ = 0.5 × *γ*_*i*_ = 0.1.

Since all simulations in Fig. 8 approximate the observed initial pandemic we need to empirically identify at least two of the three parameters *ζ, β*_*s*_, *ρ*_*s*_ to know which parameter combination most likely represents the Danish epidemic. We have previously argued for *ζ* = 0.309 in the infection model given by Eqs. (7) - (9).

If we fix *ζ* = 0.309 for the infection model in Eqs. (3) - (6) and use visual inspection we obtain a slightly lower 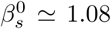 and a higher *ρ*_*s*_ ≃ 0.185 to match the initial infection peak in the reported *H*_*tot*_ data as well as the infection scale measured by May 28, 2020 (population wide antibody prevalence). However, with these parameters the later observed data requires a bit higher 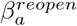.

If we instead fix 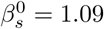 and use visual inspection we obtain *ρ*_*s*_ ≃ 0.18 and a slightly higher *ζ* ≃ 0.31 that both approximate the observed *H*_*tot*_ data and the observed May 28, 2020 population wide antibody prevalence.

Finally, if we fix *ρ*_*s*_ = 0.16 it is becomes more difficult to simultaneously fit the observed *H*_*tot*_ and antibody prevalence as of May 28, 2020. We either obtain a good seroprevalence fit and too low an initial infection wave or a good fit for the infection wave and too high a seroprevalence fit.

The above investigations with different fixed 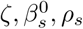 parameters underscore that even though it is always possible to identify a best MC-LS fit to *H*_*tot*_ it might not represent the actual pandemic as it misses the seroprevalence observation - and possibly other observables.

Iso-symptomatic infection diagrams could be constructed readily for most infectious diseases. With such diagrams available for the medical community, a single diagram would have crucial and quantitative information about critical parameters that define the dynamic characteristics of a given epidemic. Further, it would also at a glance make comparisons between different infectious diseases easier. Similar diagrams where critical thermodynamic properties of materials are depicted have had tremendous practical importance for the development of engineering since the early days of the industrial revolution.

### B. Estimation of mortality rates of the Danish COVID-19 epidemic

The estimated Infection Fatality Rate (IFR) as of August 31, 2020, based on the simulations that include asymptomatic cases, can be calculated by dividing the total number of deaths with the total number of recovered: 628/173, 544 = 0.0036 or ∼ 0.4%.

The official calculations of mortality of COVID-19 in Denmark in the beginning of the pandemic was based on hospitalized patients. The value was found to be 3.3% April 3, 2020, just before Easter, and 4.8% June 19, 2020, when the testing strategy had been changed for about 2 months to also include non-hospitalized individuals. However, asymptomatic cases of COVID-19 were not included in these calculations and therefore the numbers were Case Fatality Rates (CFR). Thus, according to our calculations the true mortality (Infection Fatality Rate) of COVID-19 in Denmark has been overestimated by a factor of approximately 10.

In another location of the Danish Kingdom, the Faeroe Islands (52, 484 inhabitants), testing was far more extensive from the beginning of the epidemic (109, 233 tests as of September 14, 2020) to detect and isolate *all* contacts. They have recorded 423 COVID-19 cases (0.8% including asymptomatic cases) and no deaths (both IFR and CFR) (0% - 95% c.l. 0.0 - 0.9%) [54]. Note that our estimated IFR of 0.4% in Denmark is in the middle of their confidence interval.

In Iceland (364, 260 inhabitants), a serological study estimated that 0.9% of the population had been infected with SARS-CoV-2 and that the IFR was 0.3%, which is again similar to our estimates [19].

### C. Estimation of the ℛ_0_ for the different phases of the Danish COVID-19 epidemic

ℛ_0_ can be estimated for the different phases of the epidemic using Eq. (10) if we adjust the *β*_*x*_(*t*), *x* = *i, a, s* values appropriately. For standard simulation parameters and the simulation started on February 24, 2020 with 690 incubated individuals, we estimate ℛ_0_ = 5.361 ≃ 5.4 for 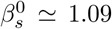 in the free pandemic until the lockdown March 16, 2020, recall Figs. 4 and 5. Using a bit lower 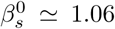 instead for the MC-LS optimized parameters yields ℛ_0_ = 5.213 ≃ 5.2. During the lockdown and the successive re-opening of the country we assume symptomatic infected individuals are effectively isolated to about 1% of the initial 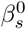 infection parameter so the majority of the infection transmission occurs due to asymptomatic and incubating individuals. We can derive 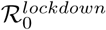 during the lockdown March 16 - April 20, 2020, and 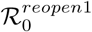 after the reopening June 8 - August 14, 2020, with infection assumptions either as in Eqs. (3) - (6) or (7) - (9). Using the standard parameters for the simulation discussed in Figs 4 and 5 based on the infection model from Eqs. (7) - (9), we have

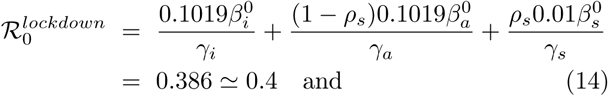

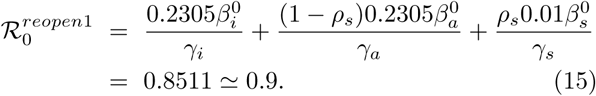

An additional Danish reopening phase occurred over the period August 14 to August 31, 2020, that most significantly allowed physical attendance at higher educational institutions and longer opening times for pubs and restaurants [53]. This last re-opening, however, was partly rolled back in the first half of September due to a significant infection increase.

An expanding epidemic requires ℛ_0_ > 1. For this to occur after the reopening of the country, the increased infection pressure must almost solely come from the asymptomatic and incubating populations as the symptomatic population is assumed to be almost completely isolated, i.e. 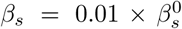. Taking this value and setting 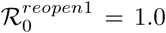 in the above equation, we can solve for the factor to multiply *β*_*a*_ and *β*_*i*_ by for the steady state reopening case and find the multiplier to be 0.2717.

From mid July to mid August 2020 the noise induced simulations generate a better approximation to the low-level steady state background infection that the observed data indicate. Neither of the deterministic simulations with parameters adjusted either by visual inspection nor by Monte Carlo Least Square optimization are able to capture the balance between the frequent micro-outbreaks and the otherwise slightly decreasing pandemic, compare Figs. 5, 6 and 7.

An approximate steady state is indicated in the following observables:

i. a fluctuating low level of hospitalized (*H*_*tot*_ ∼ 20) and
ii. a close to stationary fluctuating number of observed infected (positively PCR tested) individuals in the population at large. The infection numbers fluctuate with an average of 85/day currently based on 20,000 - 40,000 tests/day or 0.3 - 0.6% of the population/day, if we average across the re-occurring micro-outbreaks.
iii. The two contact numbers ℛ_*t*_ fluctuate around 1.0 estimated from the hospital admissions and the national testing program respectively.

We may thus view this situation as a low-level steady state for the background pandemic, with re-occurring, localized, micro-outbreaks, counteracted in part by testing and contact tracing operations. This means that Eq. (16) should modified to

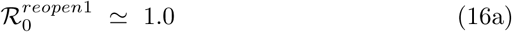

for July 15 to August 15, 2020 that indicates a quasi steady state epidemic.

## VII. ESTIMATION OF INFECTED POPULATION SIZES AND TESTING EFFICIENCIES

### A. Quasi steady state approximation for populations sizes

If we assume a steady-state pandemic situation we again have the relations given by Eq. (13). We can then use our model to estimate the sizes of the infected background populations *I*_*i*_, *I*_*a*_, and *I*_*s*_ during a quasi steady state period. It should be noted that the above mentioned steady state equation system is neutrally stable, which means that any perturbation to a solution we pick is also a solution, because the equation system is degenerate (determinant = 0). Thus, the system has infinitely many solutions all with an appropriate balance between the different infected populations and thus the resulting hospitalized patients.

Recall the hospitalization fraction *h*_*frac*_ ≃ 8.2% (Appendix B and [18]), which is the percentage of severely symptomatic infected that need hospital care, recall Eqs. (1) and (2). We can equate *I*_*s*_ × *γ*_*s*_ × *h*_*frac*_ with the average in-rate to the hospital system consisting of *H*_*tot*_ = *H* + *ICU*. Recall steady state where *dH*/*dt* +*dICU*/*dt* = 0 that can then be written out is

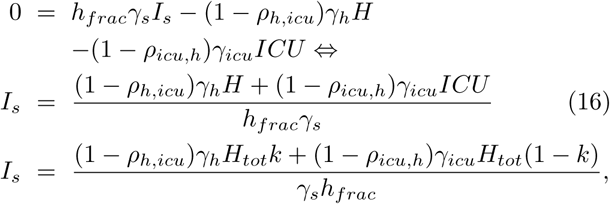

where in the last equation we have expressed *H* and *ICU* as a function of *H*_*tot*_ where *k* is defined as *H*/*H*_*tot*_. Inserting the parameter values from Table I we now have an estimate for the symptomatic population *I*_*s*_. From this we can also estimate the other infected populations as:

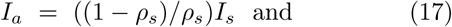

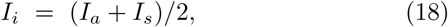

where the last relationship follows from *γ*_*a*_ = *γ*_*s*_ = 0.5*γ*_*i*_. Thus, the size of the total infected population *I*_*tot*_ is:

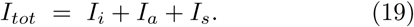

Currently PCR tests should be able to pick up SARS-CoV-2 RNA starting the last 1.5 days of the ∼ 5 day incubation period and through the ∼ 10 day period where the infected individuals are either symptomatic or asymptomatic.

Still assuming *γ*_*a*_ = *γ*_*s*_ = 0.1/day and no PCR detection after the 10 day infection period associated with *I*_*s*_ and *I*_*a*_, the detectable steady state population is approximately

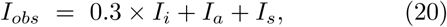

as *I*_*i*_ on average can only be identified the last 1.5 days of the 5 day incubation time; thus the factor 0.3. It should be noted, however, that RNA virus can be excreted and be picked up by PCR measurements significantly longer than the approximately 10 days we have assumed as the duration of the infection both for the symptomatic and asymptomatic cases [6].

From Eqs. (1) for the steady state case we note that

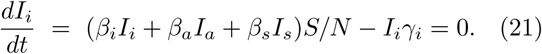

In steady state this means that the two terms should be identical; what goes into *I*_*i*_ is equal to what goes out of *I*_*i*_. Thus, we can estimate the daily rate at which newly infected are recruited from the susceptible population as

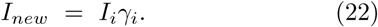

Based on hospitalization data and using Eqs. (16) to (22), we can estimate the corresponding infected and observable populations sizes as well as the daily infection rate, none of which are directly observable.

Note the linear proportionality between the numbers; e.g. 10 times higher hospital population yields 10 times higher infected populations and infection rates. Also note that these estimates may be impacted by testing and removal of identified infected individuals so their ability to further spread the infection is reduced. Finally, it should be noted that quasi steady state approximations are often successfully used even when true steady state conditions do not yet apply.

During the quasi steady state period from mid July to mid August, 2020, Denmark on average had 20 hospitalized COVID-19 patients (*H*_*tot*_) where the intensive care unit patients are approximately one fifth of every patient, *ICU* = 0.2 × *H*_*tot*_ = (1 – *k*) × *H*_*tot*_. Inserting these numbers and default values from Table I into Eqs. (16) to (20) and (22) we obtain the following population infection numbers:

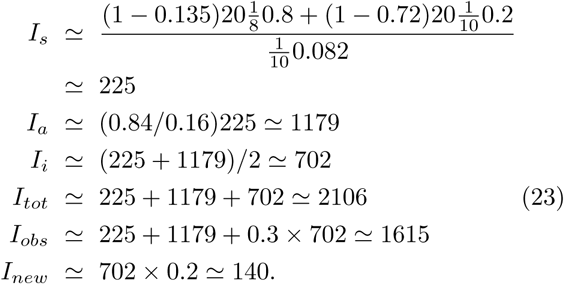

The reopening of the country August 14 - 31, 2020 unfortunately caused a new significant infection rise with an increase in the hospitalizations from early to late September. The infection rise initially had a doubling time of approximately 8 days and by the end of September, 2020 the hospital occupation largely leveled off around 110 due to a number of national initiatives including renewed information campaigns and reinstated restrictions including early closing of bars (10 pm) and at most 50 people at private events. If we assume a quasi steady state for October 1 - 20, 2020 we get from [52] the averages *H*_*tot*_ = 111, *k* = 0.85 and *T*_*d*_ = 407 that inserted in the Eqs. (16) - (22) yield:

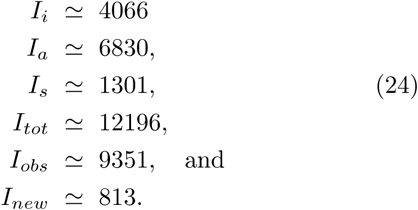

The key population numbers for different *H*_*tot*_ population sizes are tabulated in Table II (all numbers are calculated on a computer with double precision and then rounded to integer values).

**Table II:** Estimated infected population sizes and infection rates as a function of hospital population using Eqs. (16) - (22). Note the linear proportionality between the numbers; e.g., 10 times higher hospital population yields 10 times higher infected populations and infection rates. See text for details.

| Hospital & Infected Populations |  |  |  |
| --- | --- | --- | --- |
| Hospital population | Infected population | Daily infected | Observable population |
| 10 | 1053 | 70 | 807 |
| 20 | 2106 | 140 | 1615 |
| 50 | 5265 | 351 | 4036 |
| 100 | 10530 | 702 | 8073 |
| 250 | 26324 | 1755 | 20182 |
| 500 | 52649 | 3501 | 40364 |
| 1000 | 105297 | 7020 | 80728 |
| 6000 | 631784 | 42119 | 484367 |
| 15000 | 1579459 | 105297 | 1210918 |
| 30000 | 3158918 | 210595 | 2421837 |
| 60000 | 6317835 | 421189 | 4843674 |

### B. Testing efficiencies

From mid July to mid August, 2020 the nationwide voluntary testing program together with medically recommended testing and contact tracing on average identify and isolate *T*_*d*_ ∼ 85 infected/day from the contagious populations [54]. By comparing the number of identified infected *T*_*d*_ with the number observable contagious individuals *Iobs* gives a simple indicator of the testing efficiency. We may define a single day detection efficiency as the detected within a day divided with the estimated number of infected on that same day: *DT*_*abs*_ = *T*_*d*_/*I*_*obs*_ = 85/1615 = 0.0526 or approximately 5.3%. So in a theoretical situation where every individual in the country could be tested in a single day, all observably infected would be identified in one day with a total efficiency of 100%.

However, from an operational point of view a more relevant efficiency number is *DT*_*daily*_ the net daily detected and isolated compared to the daily newly infected defined *I*_*new*_. Obviously, the impact of the detection and isolation critically depends on which time point in an infected individual’s disease process it occurs. Since an explicit modeling of the infection and contact tracing processes is outside of the scope of our current study, we will only provide upper and lower bounds for the impact of the infection and contact tracing processes.

The impact would be maximal if the detection and isolation of all individuals occur during the incubation period and before the incubating infected become contagious, i.e. in the first 3.5 days of the incubation period.

Thus, we can calculate an *upper* bound of the impact of the net daily detection and isolation efficiency as 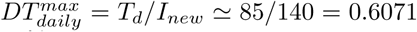 or approximately 61%.

The impact would be minimal if the detection and isolation of all individuals are asymptomatic infected occurs the very last day of their time as infectiousness. However, we need to recall that throughout this study we have assumed that almost all the symptomatic infected are detected and isolated in Denmark so part of the daily identification and isolation process involves the symptomatic infected. Since *I*_*s*_ = 225 in the steady state, we can deduce that the epidemic produces *I*_*s*_*γ*_*s*_ = 22.5 new symptomatic infected each day that are assumed detected. So 85 detections/day minus 22.5 detections/day = 62.5 detections/day are available for detection from *I*_*a*_ at their last contagious day. In our model there are 11.5 days of infectiousness for both the symptomatic and asymptomatic populations: 1.5 days of the incubation times plus 10 days of the symptomatic or asymptomatic time. If for simplicity we assume equal contagiousness every day we get that the symptomatic can only infect 1.5 days during the incubation time as we assume they are detected and isolated once they become symptomatic. The asymptomatic can infect 1.5 days during the incubation time and 9 days during the asymptomatic infection period, because they are only detected on the 10th and last day of this period.

Thus, we get a *lower* bound on the net daily detection efficiency 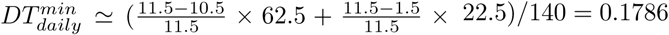 or approximately 18.0%.

We now have estimates for a lower and upper bound on the daily testing efficiency for the period July 15 and August 15, 2020 given as 18% < *DT*_*daily*_ < 61%.

As we have no detailed knowledge about the details of the infection and contact tracing processes we may use the average (center point) of the upper and the lower bound as a rough estimate for the daily testing efficiency: (18 + 61)/2 39.5 or approximately 40%. Obviously, more accurate estimates could be calculated from an explicit modeling and simulation of the involved infection and contact tracing activities.

Similarly for the period October 1 - 20, 2020 we obtain a single day detection efficiency *DT*_*abs*_ = *T*_*d*_/*I*_*obs*_ = 407/9351 = 0.0435 or approximately 4.4%. 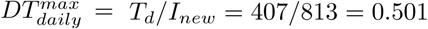 or approximately 50% while 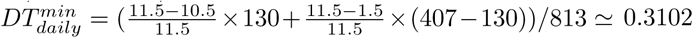 or approximately 31%.

Thus, a rough estimate for the daily testing efficiency is (50.1+31.0)/2 = 40.55 or approximately 41% that within rounding errors is the same daily testing efficiency as over the summer.

There are two important lessons to take home from the above estimates:

i. The testing, contact tracing, and isolation program in Denmark both provides a key indicator for the current geographic infection trends, but also takes part in balancing the background infection increase. This is particularly true when geographically focused efforts are undertaken. Removing and isolating on average 85 infected per day July 15 - August 15, 2020 together with behavioral restrictions had a significant impact in controlling the epidemic. Without the testing and the resulting daily removal of infected individuals the Danish pandemic would likely have been in an expansion phase during this period.
ii. The Danish testing program, which is among the most ambitious in the world, has a testing efficiency of approximately 40% for both periods investigated, while the single day infection tracing efficiency indicator is found to be 5.3% mid July to mid August 2020 and 4.4% for October 1 - 20, 2020. Many countries have both lower net daily and single day testing efficiencies as we shall see in next subsection.

### C. Hospitalized and estimated infected populations in other regions: Spain and the US

If we assume the biology of the COVID-19 pandemic is similar in Denmark, Spain and the US, and we further assume that the hospital system including *ICU*s in these countries function in a similar manner regarding treatment of COVID-19 patients, we may scale the Danish hospitalization numbers and the corresponding infected population estimates to larger populations. Obviously more detailed studies are necessary to verify - or falsify - the validity of such a comparison, but as a simplified approximation we believe such estimates provide appropriate order of magnitude estimates.

In Table II we list estimates for infected and observed population sizes based on the total hospitalized population size and the quasi steady state approximations given in Eqs. (16) to (22). For the estimates we assume the number of intensive care unit patients are approximately one fifth of every patient, thus *ICU* = 0.2 × *H*_*tot*_ (*k* = 0.8 in Eq. (16)).

Potential healthcare differences across different countries can be adjusted by adjusting the appropriate healthcare system parameters in Eq. (16).

By the end of August, 2020, Spain had about 6, 000 hospitalized with COVID-19 [39], which according to our estimates corresponds to a total infected population of about 630, 000 with about 484, 000 observable and about 42, 000 newly infected per day. In late August Spain detected and isolated a little less than 10, 000 positive COVID-19 cases/day [38]. Thus the Spanish daily single day testing efficiency indicator is approximately 10, 000/484, 000 0.0207 or 2.1%.

To estimate the net daily detection efficiency we need to approximate 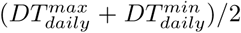.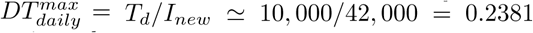 or approximately 24%. To calculate 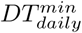 we note that *I*_*s*_ ≃ 67, 390, so *I*_*s*_*γ*_*s*_ 6, 739 new symptomatic infected are produced every day that we assume are detected. Since *T*_*d*_ 10, 000, 10, 000 6, 739 detections are thus available for individuals in *I*_*i*_ and *I*_*a*_, recall discussion in the last subsection. 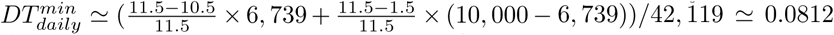 or approximately 8.1%. This gives an approximate net daily detection efficiency of (23.81 + 8.12)/2 = 15.965 or approximately 16% by the end of August 2020.

By the end of September, 2020, the US had two hospitalization peaks, both with about 60, 000 patients, one in mid April and one in late July 2020 [43], as well as two valleys, both with about 30, 000 patients, one in mid June and one mid September 2020. The corresponding estimates from Table II indicate that 30, 000 hospitalizations corresponds to about 3.16 mil. infected while 60, 000 hospitalizations corresponds to about 6.32 mil. infected. The daily detection average for the US during September 2020 was about 40, 000 [43] while the daily newly infected average was about 210, 000. Thus the single day testing efficiency indicator in the US during September was about 40, 000/2, 420, 000 0.0165 or 1.7%.

To estimate the daily detection efficiency we need to approximate 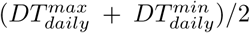.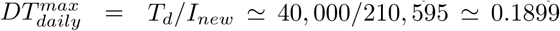 or approximately 19%. To calculate 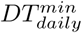 we note that *I*_*s*_ ≃ 336, 951, so *I*_*s*_*γ*_*s*_ ≃ 33, 695 new symptomatic infected are produced every day that we assume are detected. This assumption seems less appropriate with the actual numbers involved as almost all of the detections would then stem from symptomatic infected. However, to keep consistency in the estimations we keep this assumption. Since *T*_*d*_ 40, 000, 40, 000 33, 695 detections are thus available for individuals in *I*_*i*_ and *I*_*a*_, recall discussion in last subsection. 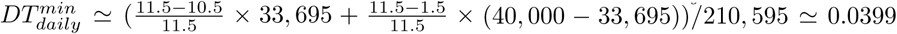 or approximately 4%. This gives an approximate net daily detection efficiency of (18.99 + 3.99)/2 = 11.4900 or approximately 11.5% during September 2020.

The above estimates from Spain and the US should only be viewed as an illustration of the method developed in Eqs. (16) - (22) applied to other countries. Healthcare differences across different countries should be implemented by adjusting the appropriate healthcare system parameters in Eq. (16) as well as our assumption about detection and isolation of symptomatic infected.

### D. Impact of the fraction of hospitalized symptomatic

Because *h*_*frac*_ is a critical part of these population estimates, we investigate in simulation the impact of both a lower *h*_*frac*_ = 0.061 and a higher *h*_*frac*_ = 0.139 using visual inspection for appropriate fits between reported *H, ICU* occupation data as well as death data and the corresponding values generated in simulation. Recall the original *h*_*frac*_ = 0.082. For details, see Appendix E.

Obviously a modified value for *h*_*frac*_ requires a significant re-adjustment of the main infection parameters. If *h*_*frac*_ increases, a larger fraction of the symptomatic infected *I*_*s*_ needs hospitalization. As the reported hospitalization volume at any given time does not change, this implies that the total number of symptomatic infected *I*_*s*_ must also be smaller to ensure correspondence between the reported and simulated data. Thus, increasing *h*_*frac*_ means a decreasing *ρ*_*s*_ so a smaller fraction of incubated *I*_*i*_ becomes symptomatic *I*_*s*_ while a larger fraction (1 - *ρ*_*s*_) becomes asymptomatic *I*_*a*_. As we know from the COVID-19 iso-symptomatic infection diagram investigation, a smaller *ρ*_*s*_ means a larger 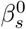 and a larger ℛ_0_ for a constant *ζ* value, that in our case is set to 0.309. As 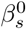 and ℛ_0_ change for the initial free pandemic so must 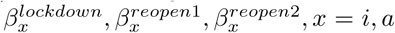, *x* = *i, a*, and their corresponding ℛ_0_ values.

Conversely, a smaller *h*_*frac*_ implies a larger *ρ*_*s*_ and a smaller *m*_*frac*_ as well as smaller *β*_*s*_ and ℛ_0_.

Nice hospital occupation and death data fits can be obtained both for *h*_*frac*_ = 0.061 and 0.139, but only for the *h*_*frac*_ = 0.139 case is it in the same simulation also possible to obtain the correct seroprevalence number of ≃ 79, 700 or ≃ 1.37% of the adult population per May 28, 2020. For *h*_*frac*_ = 0.061 the simulated seroprevalence is ≃ 96.700 and thus overshoots with a factor of 96,800/79,700 ≃ 1.21, if we at the same time require appropriate fits for *H, ICU* and deaths tolls.

For more details, see Appendix E.

## VIII. “WHAT IF?” SCENARIOS

### A. Best case scenario: Continued micro-outbreak scenarios with ℛ_0_ ≃ 1

Since April 2020 Denmark has experienced a number of localized micro-outbreaks that were contained due to isolation, contact tracing, and increasingly more extensive testing. A narrative discussion of these micro-outbreaks was given in the Introduction while in the Results Section we discuss how we model and simulate the impact of such micro-outbreaks by means of added internal noise.

Previously in Section V C we used noise to trigger injections of newly infected into the population to generate local micro-outbreaks. Local fluctuations in the behavior from time to time triggers micro-outbreaks probably in part caused by a combination of events or places where many people are gathered and super-spreaders are present. In these situations the general background dynamics is characterized by ℛ_0_ slightly smaller than 1.0 so that the background dynamics and the noise induced micro-outbreaks together generate a close to steady state infection level that corresponds to what we may call a macroscopic *effective* ℛ_0_ ≃ 1. We interpret the general background dynamics as the sum of the behavior of the general population together with the testing and contact tracing activities that result in a daily removal of infected from the epidemic, which together defines a ℛ_0_ that is smaller then one.

It should be emphasized that there is a critical difference between injecting additional infected into the general population versus changing the behavior in the general population. If the background infection is decreasing, ℛ_0_ < 1, an injection of newly infected has the same impact whether they e.g. are returning travellers that were infected elsewhere or they stem from a local micro- outbreak from within the country. In an ℛ_0_ > 1 situation both effects add equally to an already expanding pandemic.

From August 14 - 31, 2020, Denmark adjusted the re- opening of the country. The most significant changes included opening of higher educational and vocational institutions as well as longer opening hours of restaurants and bars. However, the restaurant and bar openings were partly rolled back in the first part of September together with a number of additional restrictions due to observed increase in infection levels and hospitalizations, see [53].

If, as a “best case scenario” we assume no changes in the general human behavior compared to the summer of 2020, as well as no impact by the cooler fall and winter temperatures, we obtain a development as seen in Fig. 9. We still assume recurring localized micro-outbreaks with the same frequency and amplitude as previously had, which is an optimal dynamical regime to operate in until a vaccine becomes available. Since the pandemic dynamics is operating around ℛ_0_ ∼ 1 a quasi steady state can occur for any hospital occupation level, recall Section VII A. A low hospital occupation is obviously the most desirable. Unfortunately, Denmark did not follow this scenario.

**Figure 9:**
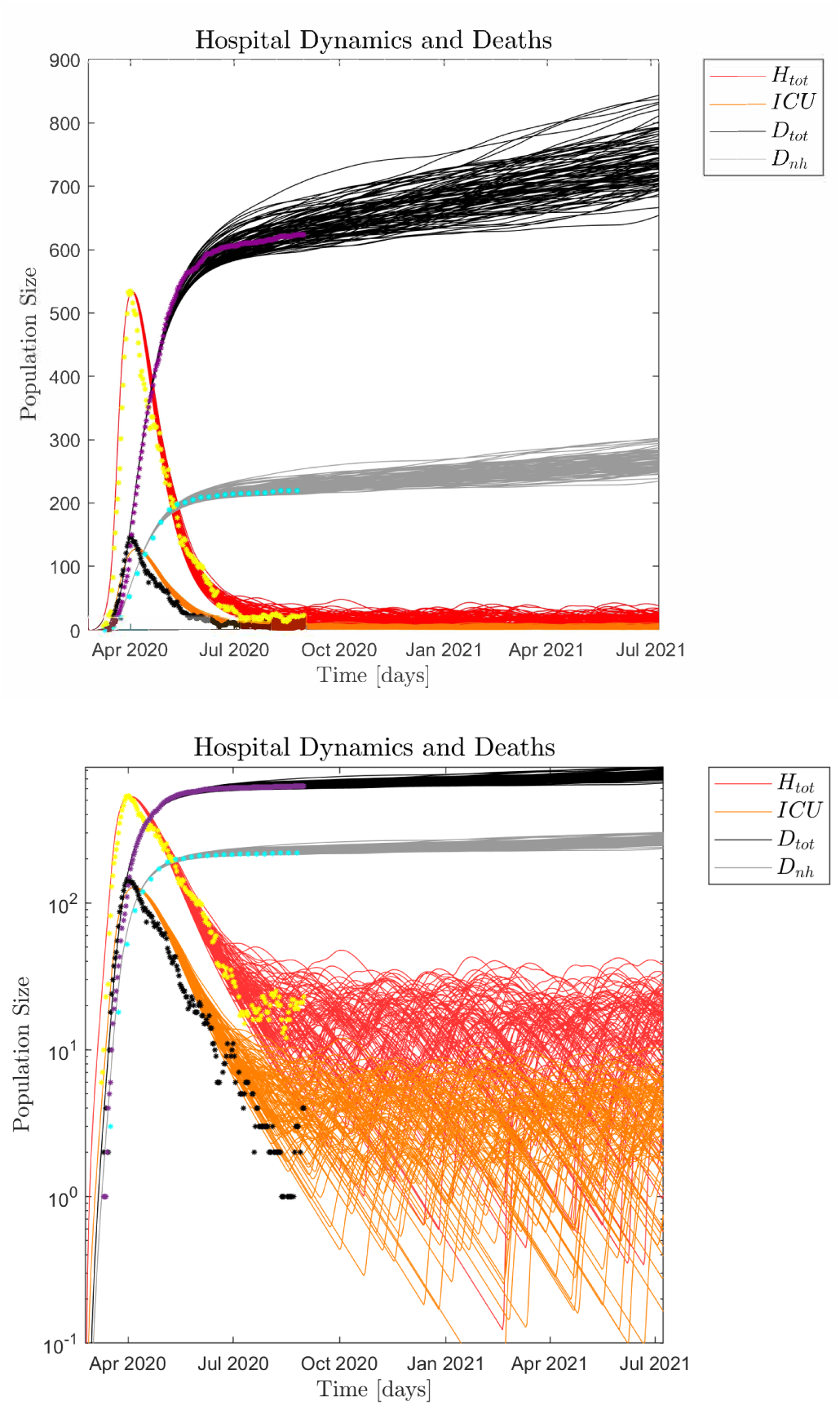
Monte Carlo simulated health care system and death toll dynamics (100 realizations) with standard parameters, recall Table 1 and Fig. 7. The pandemic is assumed to be close to steady state with successive micro-outbreaks (ℛ_0_ ∼ 1), where we assume no changes in policies and human behavior since the Summer of 2020, as well as no impact of colder fall and winter temperatures. Linear *upper panel* and logarithmic *lower panel* scales respectively. Note the continued low hospitalization and death toll. Dots represent the observed hospital and death data, lines represent the simulation data.

### B. Aggressive infection scenario following the influenza pattern

If all behavioral and policy restrictions are lifted without a universally available vaccine, a new aggressive infection wave would immediately emerge. If we further assume that we still keep the symptomatic infected effectively isolated, i.e. 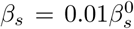, we get a scenario where the pandemic is governed by a free infection spread almost solely by the asymptomatic *I*_*a*_ and incubating *I*_*i*_ populations. Such a scenario yields, recall Eqs. (7) - (9),

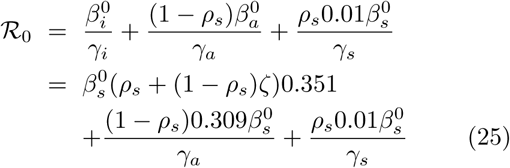

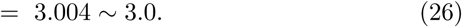

Both the initial growth in the *H*_*tot*_ observations as well as the simulations indicate that the initial pandemic February - March 2020 with ℛ_0_ ∼ 5.4 had a doubling time of ∼ 2.4 days while a pandemic with ℛ_0_ ℛ 3.0 would have a doubling time of ℛ 4.2 days. Thus, as long as we effectively isolate the symptomatic infected a worst case scenario of such an infection wave cannot be as aggressive as the initial wave we experienced early 2020. This means that future infection waves would be expected to emerge with ℛ_0_ less than 3.0 and a doubling time of more than 4.2 days.

The most vulnerable period for an aggressive Danish COVID-19 infection wave would likely be at the time the country usually experiences it’s yearly influenza epidemic, see Fig. 10.

**Figure 10:**
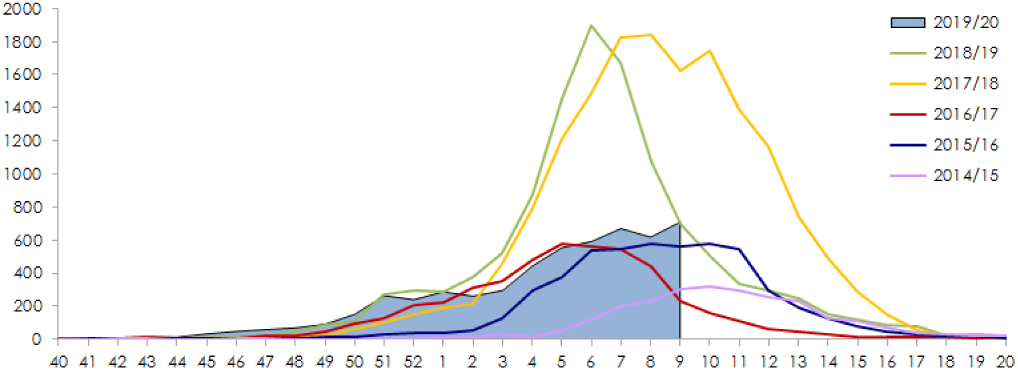
Vertical axis: number of laboratory verified influenza cases in Denmark shown for the flu seasons 2014/15 to 2019/20 until the onset of the COVID-19 pandemic. Horizontal axis: week number in a calendar year. Note the clear yearly epidemic pattern that starts in the early winter and peaks between early February and mid March. It should also be noted that the closing of Denmark due to COVID-19 March 16, 2020, stopped the influenza epidemic and actually also a smaller whooping cough epidemic, not shown in the above graph [16].

Fig. 11 shows an aggressive scenario with ℛ_0_ = 3.0 and 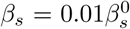 (isolated symptomatic infected) adapted to the yearly epidemic influenza pattern. To explore the full impact of such an infection wave no policy interventions are assumed, which means that incubating and asymptomatic individuals move freely to interact with and potentially infect the susceptible population. In particular, note that in particular due to the assumed limited immunological memory for the asymptotic infected (1/*ξ*_*a*_ = 60 days; decay time) the pandemic does not “burn out” as the susceptible population is continuously replenished by the recovered population.

**Figure 11:**
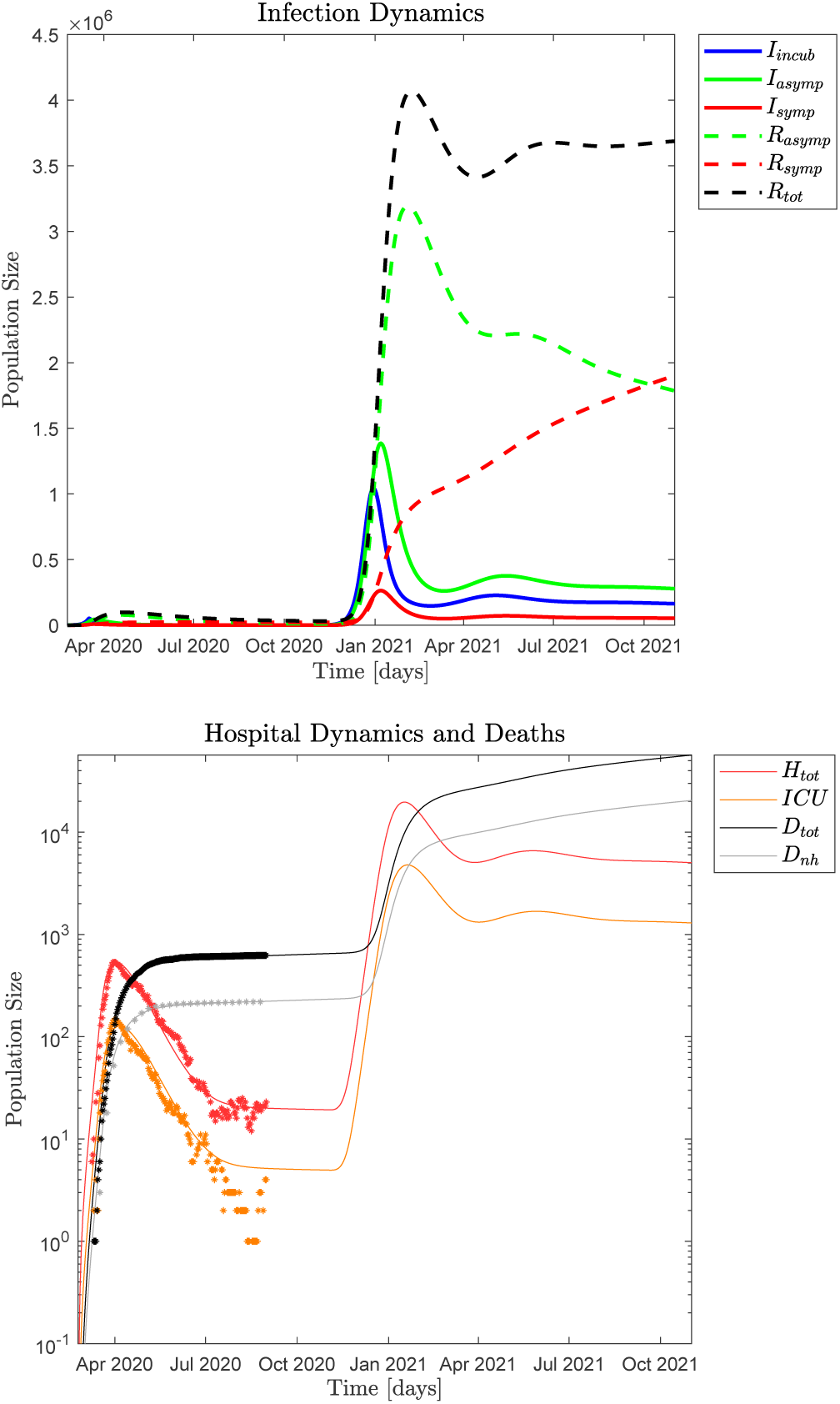
*Upper panel*: Simulated infection dynamics of infected and recovered modeled after the yearly influenza pattern using standard parameters, recall Table I and Figs. 4 and 5. This hypothetical infection wave is simulated assuming no intervention to demonstrate the full potential impact. Initially the simulation regenerates the observed hospital and death data (March - August 2020) and continues with ℛ_0_ ∼ 1 until early November 2020 when ℛ_0_ in the simulation in- creases to 3.0 recall Eq. (26). *Lower panel*: Simulated resulting total hospital and *ICU* occupations, as well as total deaths and non-hospital deaths in logarithmic scale. Dots represent the observed hospital occupation and death data, lines represent the simulation data. After a year (November 1, 2021) mainly due to the short immune memory of the asymptomatic infected (decay-time ∼ 60 days), a close to steady state situation of the pandemic would be reached with *H*_*tot*_ ≃ 5, 027 out of which *ICU* ≃ 1, 295. We may view such a scenario as a “soft” version of the free COVID-19 pandemic explored in Fig 12.

Without intervention the pandemic peaks for *I*_*i*_ December 30, 2020 and January 7, 2021 for *I*_*a*_ and *I*_*s*_, and by February 7, 2021, appoximately 4, 071, 000 people (∼ 70% of the population) would have been infected and recovered from the pandemic. The simulated hospital impact peaks around January 17, 2021 with ∼ 19, 680 hospitalized and January 20, 2021 with ≃ 4, 786 in *ICU*s which would completely overwhelm the Danish health- care system. It is estimated that the Danish *ICU* capacity is about 1, 000 beds and the total hospital capacity is about 15, 000 beds. A year after the onset of this infection wave (November 1, 2021) about 56, 690 people would have died from the SARS-CoV-2 virus, which is about the same as the total number of deaths in Denmark in the year 2019: 53, 958.

### C. Free pandemic: Worst case scenario

“What would have happened if no policy interventions were imposed at the beginning of the pandemic?” This is a question many people have asked us. In Fig. 12 data are generated in simulation using standard the parameters also used in Figs 4 and 5, but assuming no behavioral modifications or policies imposed at the onset of the pandemic, thus ℛ_0_ ≃ 5.36. This is a hypothetical scenario where the pandemic rages freely.

**Figure 12:**
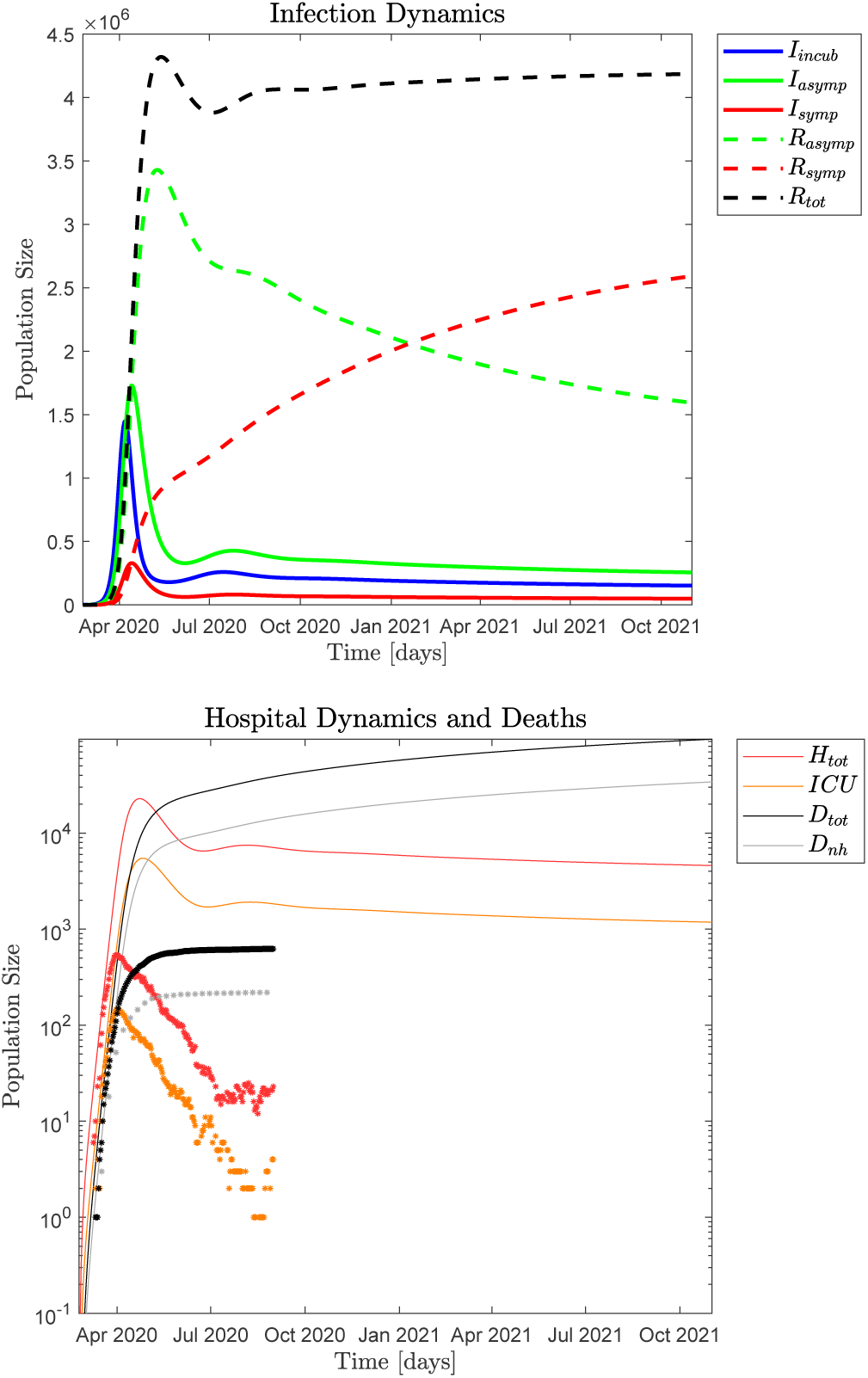
*Upper panel*: The simulated dynamics of the infected and recovered for a hypothetical free COVID-19 pandemic in Denmark that started February 24, 2020 and runs until November 1, 2021. This hypothetical pandemic would have peaked April 7, 2020 with *I*_*i*_ ≃ 1, 451, 000 and peaked April 13 with *I*_*a*_ ≃ 1, 729, 000 and *I*_*s*_ ≃ 329, 400 infected individuals. By May 13, 2020, the recovered population would have peaked with more than 4.32 mil. or about 74 % of the population (*R*_*tot*_ ≃ 4, 320, 000). *Lower panel*: Simulated and reported health care system occupation as well as death toll data (logarithmic scale). Dots represent the observed hospital and death data, lines represent the simulation data. The hypothetical death toll of 64, 960 is clearly underestimated as we use the health care model from Eqs. (2) that in this simulation is assumed to be able to scale with the pandemic, which it clearly cannot in real life. The hospitalization occupation peaks April 23, 2020, with *H*_*tot*_ ≃ 22, 850 patients and the intensive care units peaks April 26 with *ICU* ≃ 5, 472. The Danish healthcare systems would be completely overwhelmed in such a scenario and we would expect an increased mortality rate because of that.

The pandemic does not “burn out” in this scenario either as the susceptible population is continuously replenished by the recovered population, recall discussion in Fig. 11. This means that a relatively stable hospital population of about *H*_*tot*_ ≃ 6, 500 by October 2020 would slowly decrease to ≃ 4, 500 in October 2021. A conservative estimate of the expected excess deaths from such a free pandemic is ≃ 64, 960 after one year, by February 24, 2021, which is more than all the deaths in Denmark in 2019 (53, 958).

## IX. DISCUSSION

The goal of this work is to provide a theoretical platform to help understand the details of the Danish COVID-19 epidemic that can hopefully also provide useful in-sights about the nature of the pandemic in other countries and regions. Our models and simulations can easily be adapted to other regions or countries where hospital, *ICU* and death data are available.

We have chosen a simple macroscopic description and used a differential equation based simulation of the pandemic and health care system dynamics that seems appropriate for most of the issues we seek to address, although a microscopic agent based simulation scheme would have been more appropriate e.g. to capture the dynamics of the observed micro-outbreaks. Our approach is also limited in two other aspects. We neither have a geographic representation of the pandemic nor do we have an age disaggregated population although reported data clearly show features caused both by geography and age differences in the population.

However, by adding noise to our model and conducting Monte Carlo simulations of large ensembles of the system we can to some extent compensate for the shortcomings caused by the lack of geography representations and the occurrence of localized micro-outbreaks. This approach is further supported because Denmark is a small country with a relatively high mobility and population mixing. The missing age distribution does not cause serious problems for understanding the initial epidemic dynamics, but for the later micro-outbreaks we in principle need to know the nature of their age distribution as this significantly impacts the health care system. We have not implemented such parameter changes in our simulations. All parameters are kept constant over time in each of our simulations except the externally imposed lockdown and reopening policies/behaviors. By adjusting the amplitude and frequency of the added noise we can adjust the expected impact on the health care system from such micro-outbreaks.

The micro-outbreaks since late July 2020 began in a slaughterhouse in the city of Ringsted. Then came the outbreaks in the three largest cities in Denmark: first in Århus and then around September 1, 2020 in Odense and Copenhagen. These outbreaks were driven by young people aged 20-29 years and immigrants from Middle East and Somalia who live in rather closed communities.The outbreaks spread to other groups but fewer old and vulnerable persons became infected compared to the epidemic in the spring. The number of patients hospitalized, in particular patients treated in *ICU*s and the number of deaths, remained very low in the fall period compared to the spring epidemic although it was still the old people who dominated the hospitalizations and deaths. In each of the micro-outbreaks nonphamacological measures were re-introduced and the ℛ_*t*_ of each micro-outbreak went from above 1.5 to approximately 0.8 within a month, which probably is to be expected considering the epidemic dynamics of COVID-19.

The testing capacity during these outbreaks went up to nearly 1% of the whole Danish population each day and by October 7, 43% of the population had been tested for SARS-CoV-2. That means that many persons that contacted SARS-CoV-2 positive persons including asymptomatic persons were detected. Therefore of the 9, 623 SARS-CoV-2 positive persons in the fall outbreak only 34 (0.37%) died, whereas 434 (4.8%) of the 8, 851 SARS-CoV-2 positive patients who were hospitalized during the spring epidemic died. The reduced mortality is similar to a case-fatality rate of 0.4% in a recent report from Hong Kong [27].The early high rate mainly reflects that only patients who were hospitalized were tested for SARS-CoV-2 in the spring epidemic. Therefore, the true mortality of the spring epidemic was calculated too high. This is important to realize, because a restricted testing capacity may mislead the health-care system, the population and the precautions taken to manage the epidemic, e.g. based on the calculated ℛ_0_ and ℛ_*t*_. The burden on the hospitals and ICUs was much lower in the outbreaks in the fall. The spring/fall ratio of the maximum number of normal hospital unit COVID-19 patients was 535/147 while the corresponding ratio for *ICU* patients was 153/20. The improved treatment of COVID-19 patients (with e.g. Remdesevir, dexamethasone, and prophylaxis and treatment of bleeding disorders, [5][12]) is probably the reason for the proportionally lower number of hospitalized patients who in the fall were treated in ICUs or died.

The estimated percentage of symptomatic infected that needs hospital care *h*_*frac*_ is a critical parameter as it scales the connection between the main observables, the time series for *H, ICU* occupations, and the death toll, and the infection dynamics. We have used an age aggregated estimate for *h*_*frac*_ = 8.2% based on the age distribution of health care needs from [18]. With an increase of *h*_*frac*_ to 13.9% the simulation still corresponds reasonably well to the observed data we have access to, while a decrease in *h*_*frac*_ to 6.1% does not allow the simulation to both satisfy the time series of our main observables and the measured seroprevalence of May 28, 2020.

For our hospital model we have also chosen simplicity over including more details that potentially could help capture the health care dynamics in greater detail. The simple hospital model is nonetheless able to reproduce both the historical hospital and death data quite well, but it has difficulties reproducing some of the reported data details. For example, both the reported *H*_*tot*_ and *ICU* occupation time series have sharper peaks at the onset of the pandemic than the simulated peaks. Also to enable the simulated *ICU* data to follow the reported time series data in a reasonable manner we need to use an average *ICU* occupation time 1/*γ*_*icu*_ = 10 days, which is shorter than the reported average occupation time, although it should also be noted that a more recent improved treatment of COVID-19 patients means that fewer have a severe course [5] [12].

Requiring our healthcare model to reproduce the historical hospital *H*_*tot*_ and *ICU* data, the simulation underestimates the total accumulated number of hospital patients *H*_*tot*_ by 7-14% and it also underestimates the accumulated number of *ICU* patients. Thus, our simple health care model has difficulties simultaneously matching both the time series data (daily hospital occupation numbers) and the accumulated data (total number of hospitalized patients).

There is an ongoing discussion of the best value for average period of infectiousness both for the symptomatic and asymptomatic populations, where [6] suggests 1/*γ*_*s*_ = 1/*γ*_*a*_ = 10 days while [28] suggests 7 days as a better estimate. Lowering the infectious period 1/*γ*_*s*_ and 1/*γ*_*a*_ both for the symptomatic and the asymptomatic populations from 10 to 8 days in our simulation makes it difficult to reproduce the total hospital occupation numbers (too fast a decline of simulation numbers after lockdown), while for the simulated death numbers from the hospitals (too fast a rise). However, shorter 1/*γ*_*s*_ and 1/*γ*_*a*_ values make it easier to reproduce the initial relatively sharp peak for the *ICU* occupation number. To compensate for the lower 1/*γ*_*a*_ and 1/*γ*_*s*_ values the *β* values must increase a bit. With lower infection times minor adjustments are needed for the parameters associated with hospitals, *ICU*s, and death tolls.

We can use Monte Carlo (MC) optimization of the parameters where we minimize the Least Square (LS) difference between the reported and the simulated time series of the total hospital occupation (*H* + *ICU*). Despite the minor mismatches between what the MC-LS parameter optimization yields and what visual parameter inspection yields, the MC-LS capability enables us to do large scale explorations of the parameter impacts.

Using a steady state approximation for the background infection that is indicated by approximately constant low values in our main observables, i.e. *H, ICU* and death data, enables us to make simple analytical estimates of the sizes of the corresponding infected populations *I*_*i*_, *I*_*a*_ and *I*_*s*_. These analytical estimates are robust to most parameter perturbations and they are confirmed by numerical simulations using steady state conditions. This approach and the corresponding infected population estimates should give more accurate predictions of the actual sizes of the infected populations than direct infection testing of the daily scale currently being done.

Another general finding coming out of this study is the tight relationship that exists between *β*_*s*_, ℛ_0_, *ρ*_*s*_ and *ζ*, which can be represented by an iso-symptomatic- infection diagram, see Section VI A, that is made possible by using MC-LS optimization for each *β*_*s*_, ℛ_0_, *ρ*_*s*_ and *ζ* parameter combination. We propose such diagrams could readily be constructed for most infectious diseases, so that one diagram would hold crucial and comparable quantitative information about critical parameters that define the dynamic characteristics of a pandemic.

## X. CONCLUSION

Our computational platform explores the connection between the dynamics of the viral epidemic, the national health care system and the imposed policies.

Based on reported seroprevalence data [54] we can estimate the relative frequency between symptomatic and asymptomatic infected to be 16% and 84% respectively. We estimate ℛ_0_ ∼ 5.4 for the initial free pandemic, ℛ_0_ ∼ 0.4 for the lockdown period and ℛ_0_ ∼ 0.8 *–* 1 for most of the successive reopening periods.

For the immediate future, as the northern hemisphere prepares for the upcoming winter and potential new infection waves, it should be noted that over the summer Denmark has been operating with *R*_0_ ∼ 1 from June through August 2020. The estimated infected population sizes for the quasi steady state period mid July to mid August, 2020 are *I*_*i*_ ≃ 702; *I*_*a*_ ≃ 1, 179; *I*_*s*_ ≃ 225; *I*_*tot*_ ≃ 2, 106; and *I*_*obs*_ ≃ 1, 615, while the daily infection rate is *I*_*new*_ ≃ 140.

The daily net testing efficiency is estimated to be approximately 40% for the periods July 15 - August 15, 2020 and October 1 - 20, 2020. The single day identification efficiency indicator in these period, defined as the number of positively identified infected over the estimated observable infected population the same day, is about 5%.

Since the symptomatic infected are effectively isolated from the population, and if behavioral policies are kept in place and observed, future COVID-19 infection waves fortunately would not be nearly as aggressive as the first wave assuming the viral biology does not change significantly due to mutations. We estimate that a new infection wave would have a lower bound with doubling time of more than 4.2 days and less than ℛ_0_ ∼ 3.0 compared to the initial wave with a doubling time of about 2.4 days and ℛ_0_ ∼ 5.4.

We believe our simulation platform is suitable for exploring forecasting scenarios for the upcoming winter and beyond or until a vaccine hopefully becomes available. Our results indicate that it is possible to more precisely than previously calculate the burden on the hospitals and *ICU*s of new COVID-19 infection waves or a new pandemic. Because of that, our simulation could hopefully also help minimize the impact on hospital treatment of non-COVID-19 patients.

Importantly we have shown, that the mortality of COVID-19 in Denmark is only ∼ 0.4% like in Iceland and the Faeroe Islands. Although a mortality rate of~0.4% may seem small, our simulation indicates that had Denmark not adopted any behavioral measures to counteract the Danish pandemic the death toll would be ≃ 64, 960 by February 24, 2021 one year into the pandemic. That surpasses the total number of deaths (53, 958) in Den- mark in the year 2019. This simulation based COVID-19 death estimate is conservative as we have assumed a fully functional healthcare system, which is unrealistic as the free pandemic in such a scenario would have resulted in a hospital *H*_*tot*_ and *ICU* occupation peaks in late April, 2020 that would have completely overwhelmed the Danish health care system with ≃ 22, 850 hospitalized and ≃ 5, 472 in *ICU*s. Per January 1, 2021 there would still be ≃ 5, 860 hospitalized and 1, 500 in the *ICU*s in a free COVID-19 pandemic according to such a simulation scenario.

Clearly Denmark’s adopted behavioral modifications, lockdown, measured reopening, testing, and contact tracing procedures, have been highly successful in mitigating the epidemic from the onset through August 2020.

## Data Availability

All data used are publicly available data that are clearly referenced in the manuscript

## Acknowledgment

We are grateful for the general guidance of John Erik Hansen regarding all epidemic matters in the early stages of this study as well as a final critical review of the manuscript. We are also grateful for the enlightening conversations with Gitte Kronborg regarding the COVID-19 situation at Danish hospitals in the early stages of the epidemic. Oana Ciofu and Asbjørn Dahl are acknowledged for their critical review of the manuscript and we thank Hans Ziock for his in depth critical reading, corrections, and constructive suggestions throughout the last versions of the manuscript. All remaining errors in the manuscript are the responsibility of the authors.

## Appendix A Statens Serum Institut data

COVID-19 data from Statens Serum Institut is plotted in Fig. 13 (data accessed per August 31, 2020) See caption for details.

**Figure 13:**
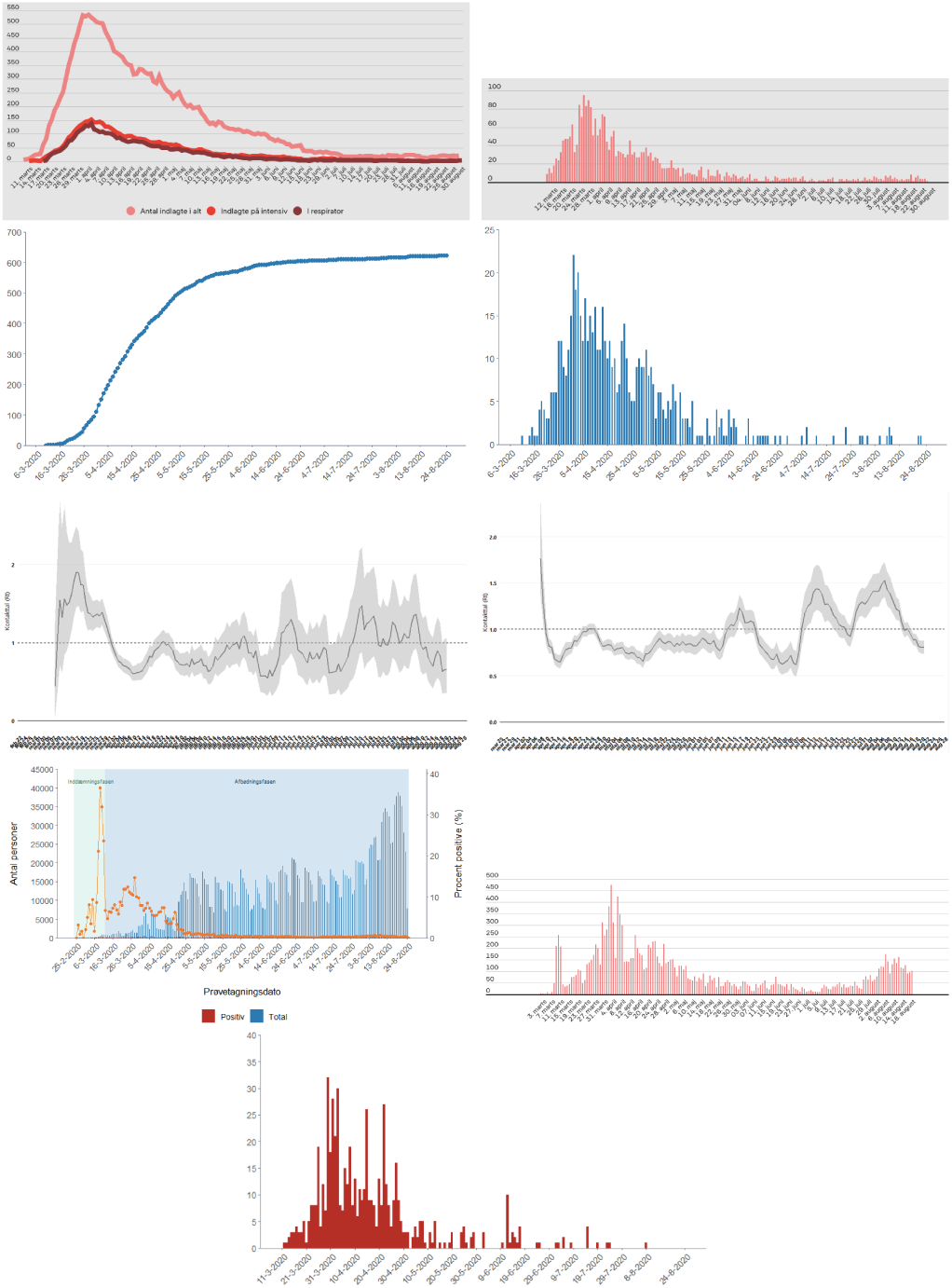
*Upper panel*: (1) Daily national COVID-19 hospital occupations and (2) daily admissions. Note the “shoulder” in the admission data around mid April 2020 indicated by a nearly constant admission level for a couple of weeks that at the time breaks a close to exponential decrease in the pattern of the daily admissions. *Upper middle panel*: (3) accumulated daily deaths and (4) daily deaths. *Lower middle panel*: (5) ℛ_*t*_ calculated from the daily hospital admission data and (6) ℛ_*t*_ calculated from the nationwide testing program. Note that in the period April 1 - August 31, 2020, one can distinguish five distinct peaks in ℛ_*t*_ both measured from the hospital data and from the PCR count. Before about April 1, 2020, the epidemic was dominated by the initial dramatic pandemic growth and successive lockdown of the country making it impossible to identify earlier distinct microscopic events in the data. *Lower, lower middle panel*: (7) daily number of PCR tested nationwide, as well as positively tested and (8) the daily number of positively tested shown at a different scale. Note how the capacity of the national testing program had been stable at around 20,000 tests/day since early May, see image (7), and that the capacity was expanded in August as a result of a local micro-outbreak that started in the city of Aarhus that grew and started to spread to other regions. The national capacity is planned to be expanded to 50-60,000 tests/day by mid fall 2020. *Lowest panel*: (9) the daily number of positive PCR tests at home care centers. Note how increased caution and infection prevention policies over time have significantly decreased the infection level in the older part of the population reflected in the observed number of infected at home care centers.

It should be noted that SSI has modified minor details in data multiple times during the period February through August 2020. However, these changes have not had any measurable impact on our investigations or conclusions.

## Appendix B Fraction of hospitalized symptomatic infected

The current age distribution of the population in Denmark is given by Danmarks Statistik (Statistics Denmark) [11] and by combining it with the age dependent hospitalization fractions given in Ferguson et al.(2020), p5, table 1 [18] yields:

| Age distribution of hospitalization |  |
| --- | --- |
| A0019=1.30/5.83 | $H_{frac0019}=(0.1+0.3)/2$ |
| A2039=1.47/5.83 | $H_{frac2039}=(1.2+3.2)/2$ |
| A4059=1.56/5.83 | $H_{frac4059}=(4.9+10.2)/2$ |
| A6079=1.23/5.83 | $H_{frac6079}=(16.6+24.3)/2$ |
| A80up=0.27/5.83 | $H_{frac80up}=27.3$ |

From these age weighted hospitalization fractions, *h*_*frac*_ is obtained as

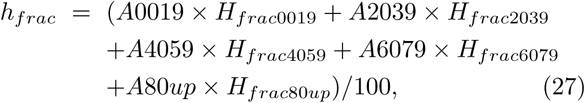

which yields *h*_*frac*_ = 0.0820.

Obviously, the value of *h*_*frac*_ is an important parameter as it scales the epidemic because it determines how many symptomatic infected *I*_*s*_ enter the hospital system, which is the empirical data we use to adjust our simulation. A smaller *h*_*frac*_ implies a larger scale epidemic with higher total number of infected individuals nation- wide while a larger *h*_*frac*_ implies a smaller scale epidemic. Note that the overall scale of the epidemic is in part restricted by the empirical hospital and death data and in part by the empirical data from the population wide serological antibody count. A detailed discussion of the impact of the size of *h*_*frac*_ is given in Appendix E.

## Appendix C Monte Carlo Least Square optimization

The optimization algorithm includes a Monte Carlo method to minimize the sum of squared differences between the Danish hospitalization data and corresponding data generated in simulation, which is the sum of the state variables *H* and *ICU* in the model, recall equations (1) and (2). The Least Squares (LS) difference is defined as follows:

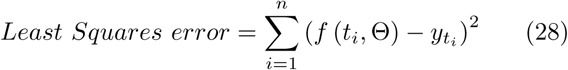

where 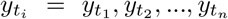 is the observed data and *f* (*t*_*i*_, Θ) is the corresponding solution of the simulation with a given parameter set Θ = (*θ*_1_, *θ*_2_, …, *θ*_*m*_). A pseudo description of the algorithm is as follows:

i. Generate random numbers within a given domain and choose these numbers as values for selected parameters,
ii. Run a simulation with these parameter settings,
iii. Calculate the LS difference between reported and simulated data with chosen parameters,
iv. If LS error is less than the former LS error then save the parameter values,
v. Repeat a predefined number of times - or until a certain small LS error value is reached.

Figure 14 illustrates the Least Squares error for a standard run with *ρ*_*s*_ = 0.16 and *ζ* = 0.309 only sampling on values of 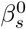 from a uniform distribution. Note the minimum of the LS error for 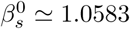.

**Figure 14:**
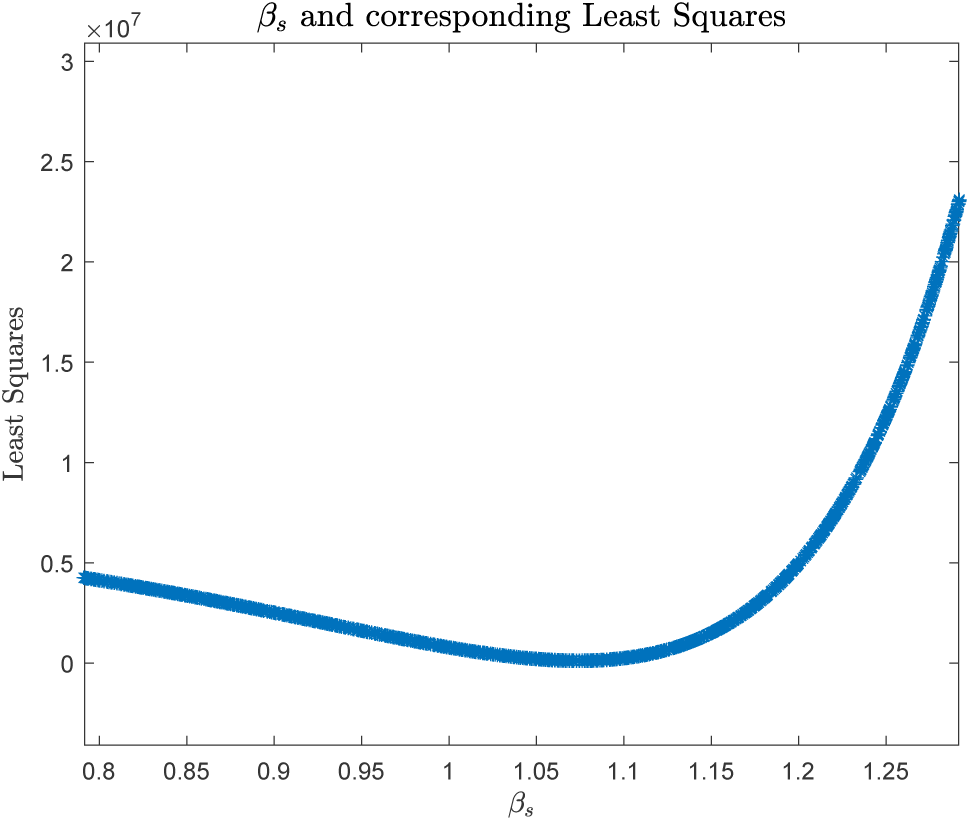
Least Squares values produced by random selected 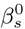 -values with a minimum for 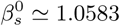. Recall parameter discussion in Monte Carlo Least Squares section.

## Appendix D Derivation of ℛ_0_

The basic reproduction number, ℛ_0_, can be calculated from an epidemic compartment model using the Next-Generation Matrix Method [10]. Our epidemic compartment model given in Eqs. (1) consists of five compartments (*S, I*_*i*_, *I*_*a*_, *I*_*s*_, *R*) with three (*I*_*i*_, *I*_*a*_, *I*_*s*_) infected populations. Letting *S* = *N* (total population) and only including the infected individuals Eqs. (1) reduce to

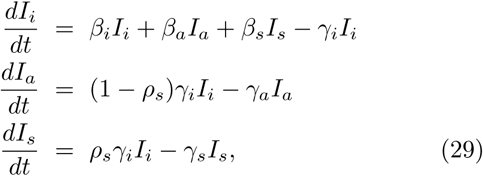

where we have ignored the hospital dynamics. We may now write Eqs (29) in matrix form as

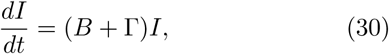

where the *B* matrix express the epidemic *transmissions* while the Γ matrix express the *transitions* between the infected populations each defined as

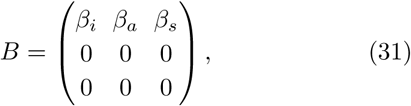

and

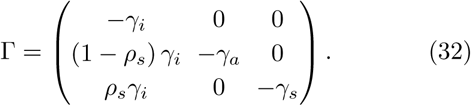

ℛ_0_ can now be defined as [10]

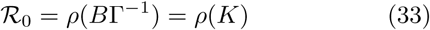

where *ρ*(*K*) defines the dominating eigenvalue of the resulting *K* matrix.

The inverse of Γ is calculated by 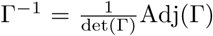. The determinant of Γ is

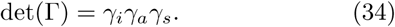

By first transposing Γ we may calculate the adjunct of Γ by first transposing, then finding the matrix of the minor and then calculating the matrix of cofactors.

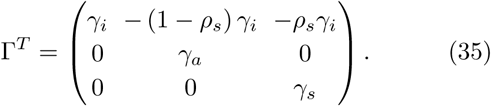

The matrix of minors is given as

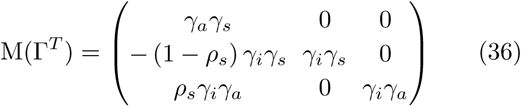

Applying the matrix of cofactors to (36) to obtain the adjunct of Γ

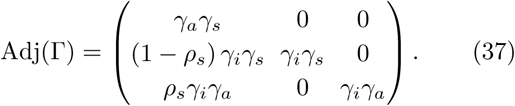

This implies

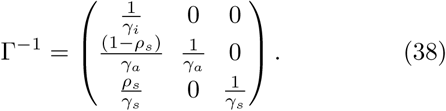

By matrix multiplication of *B* and Γ^−1^ we get

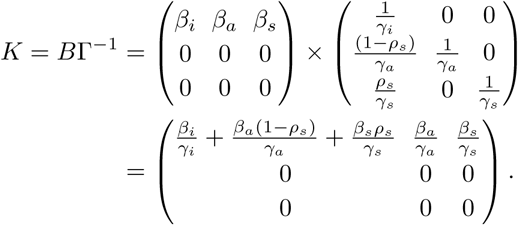

Since *K* = *B*Γ^−1^ is an upper triangular matrix the (only) eigenvalue can be read off the diagonal and the basic reproduction number is

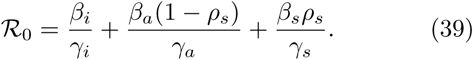

## Appendix E Impact of size of *h*_*frac*_

Here we address the impact of changing the size of *h*_*frac*_. If *h*_*frac*_ increases a larger fraction of the symptomatic infected *I*_*s*_ needs hospitalization. As the reported hospitalization volume at any given time does not change, this implies that the total number of symptomatic infected *I*_*s*_ must also be smaller so as to ensure correspondence between the reported and simulated data. Thus, increasing *h*_*frac*_ means a decreasing *ρ*_*s*_ so a smaller fraction of incubated *I*_*i*_ becomes symptomatic *I*_*s*_ while a larger fraction (1 - *ρ*_*s*_) becomes asymptomatic *I*_*a*_. As we know from the COVID-19 iso-symptomatic infection diagram investigation, a smaller *ρ*_*s*_ means a larger 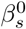 and a larger ℛ_0_ for a constant *ζ* value, that in our case is set to 0.309. As 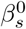 and ℛ_0_ change for the initial free pandemic so must 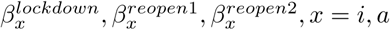, *x* = *i, a* and their corresponding ℛ_0_ values.

We test two concrete *h*_*frac*_ values away from the standard *h*_*frac*_ = 8.2%:

i. *h*_*frac*_ = 13.9%, a ∼ 70% increase, see Fig. 15.

**Figure 15:**
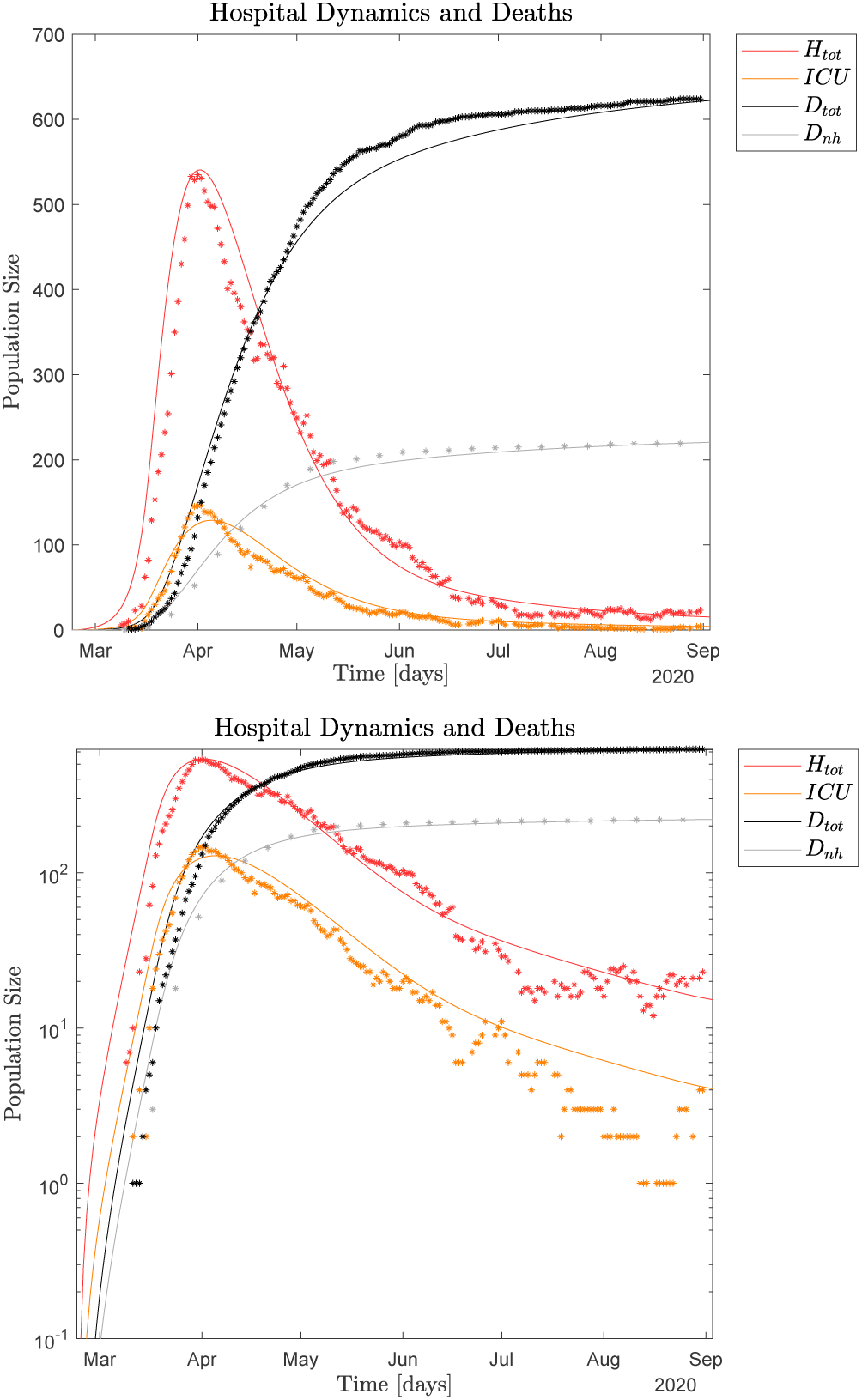
*Upper panel*: Reported and simulated hospital *H*_*tot*_, *ICU* and death data for *h*_*frac*_ = 13.9% linear scale. Simulated seroprevalence matches with the observed May 28, 2020 value. Necessary parameter adjustments compared to standard simulation, recall Table I. Epidemic parameters: 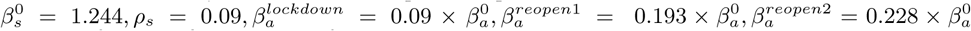 As expected, the fraction of terminally ill non-hospital patients is as expected significantly increased *m*_*frac*_ = 0.0131, while the hospital system parameters are only slightly changed: *ρ*_*h,r*_ = 0.78, *ρ*_*h,icu*_ = 0.135, *ρ*_*h,d*_ = 0.085. The rest of the simulation parameters are the standard values given in Table I. *Lower panel*: Same reported and simulated observables but logarithmic scale. Dots represent the observed hospital and death data, lines represent the simulation data.
ii. *h*_*frac*_ = 6.1%, a ∼ 25% decrease, see Fig. 16.

**Figure 16:**
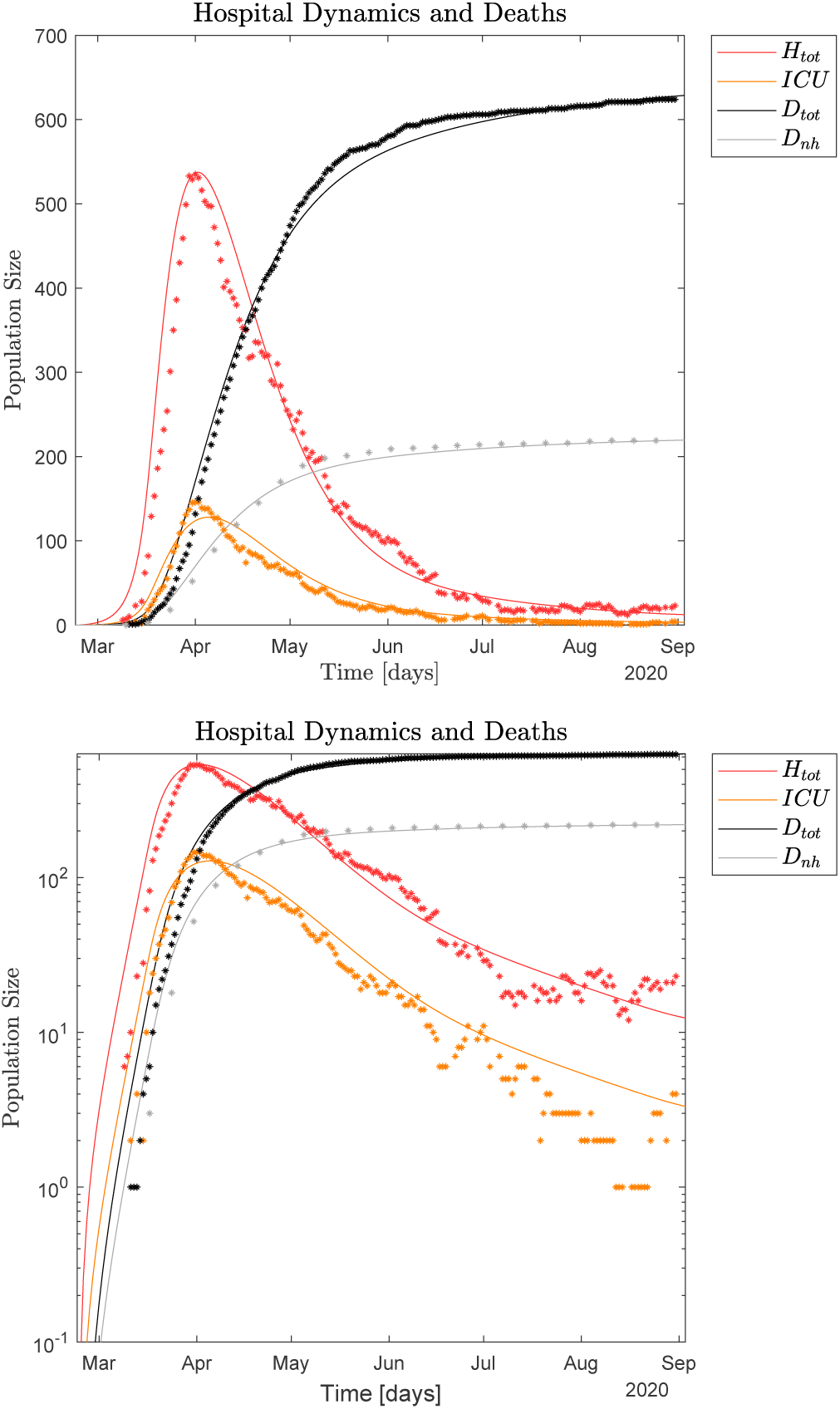
*Upper panel*: Reported and simulated hospital *H*_*tot*_, *ICU* and death data for *h*_*frac*_ = 6.1% linear scale. Simulated seroprevalence of ∼ 96, 400 exceeds observation of ∼ 79, 700 May 28, 2020. Necessary parameter adjustments compared to standard simulation, recall Table I. Epidemic parameters: 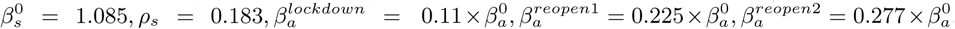 No adjustments of the health care parameters are necessary, but as expected *m*_*frac*_ = 0.0058 is significantly lower because *I*_*s*_ becomes larger due to the smaller *h*_*frac*_. *Lower panel*: Same reported and simulated observables but logarithmic scale. Dots represent the observed hospital and death data, lines represent the simulation data.

Figs. 15 and 16 should be compared with Figs. 4, 5 and 6.

Although both simulation experiments in Figs. 15 and 16 generate nice correspondence with the observed primary data, only *h*_*frac*_ = 13.9% is simultaneously able to satisfy the reported seroprevalence of May 28, 2020. This means that 0.082 ≤ *h*_*frac*_ ≤ 0.139 can satisfy all observed data with parameter adjustments, while *h*_*frac*_ = 6.1% cannot. See more details in the figure captions.

